# Defining ocular *Chlamydia trachomatis* infection prevalence thresholds for the trachoma elimination endgame

**DOI:** 10.64898/2026.11.02.26366918

**Authors:** Anna Borlase, Jacob A. Cohen, Everlyn Kamau, Rosemary Pearson-Clarke, Kristen K. Renneker, Fentahun Tadesse Akale, Paul Akampurira, Abdou Amza, Sue Chen Apadinuwe, Solomon Aragie, Sylvie Araka, Zebene Ayele, Getahun Ayenew, Mohamed S. Bah, Robin L. Bailey, Ana Bakhtiari, Stephen Begumisa, Tesfahun Bishaw, Yak Yak Bol, Lochebe Boniface, Sarah Boyd, E. Kelly Callahan, David Chaima, Benard Chieng, Ambahun Chernet, Adisu Abebe Dawed, Michael Dejene, Dereje Duguma, Belgesa E. Elshafie, Samuel Eshitemi Omukuba, Mawo Fall, Mohammed Feyisso Shaka, Katherine Gass, Teshome Gebre, Tania A. Gonzalez, Henry Emisiko, Emma M. Harding-Esch, PJ Hooper, Kimberly A. Jensen, Cristina Jimenez, Boubacar Kadri, John M. Kaldor, Khumbo Kalua, Henry M. Kanyi, Mabula Kasubi, Fikreab Kebede, Jeremy D. Keenan, Amir B. Kello, Richelle W. Kihoro, Namasiku S. Kunda, Stephen B Lambert, Sara Lavinia Brair, Kathleen Lynch, Benjamin Marfo, Michael Peter Masika, Stephen Gabriel Mbwambo, Harran Mkocha, Tamimo Momade, Caleb Mpyet, Kangwa I. M. Muma, Constity Mwale, Clarer Jones, Beido Nassirou, Susana Vaz Nery, Jeremiah Ngondi, Sammy M. Njenga, Stephen Ohidor, Catherine Oldenburg, Nicholas Olobio, Rapheal Opon, William E Oswald, Fatai Oyediran, Martha Idalí Saboyá Díaz, Angelia M. Sanders, Eshetu Sata, Fikre Seife, Laura G Senyonjo, Esther Andia Shiraho, Henis Mior Sitoe, Hiwot Solomon, Jesse Speer, Zerihun Tadesse Gebreselassie, Sandra Liliana Talero Castro, Emma Michelle Taylor, Teyil Wamyil-Mshelia, Titus Watitu Kimani, Sheila K. West, Dionna M. Wittberg, Dorothy Yeboah-Manu, Mulat Zerihun, Taye Zeru, Thomas M. Lietman, Scott D. Nash, Benjamin F. Arnold, Anthony W. Solomon

**Affiliations:** Department of Biology, University of Oxford, Oxford, UK; Francis I. Proctor Foundation, University of California San Francisco, San Francisco, CA, USA; International Trachoma Initiative, Task Force for Global Health, Atlanta, GA, USA; Sightsavers, Addis Ababa, Ethiopia; Vector-Borne and NTDs Molecular Laboratory, Kampala, Uganda; Programme National de Lutte Contre la Cecité, Niamey, Niger; Ministry of Health, Yaren District, Republic of Naoero; The Carter Center, Addis Ababa, Ethiopia; Eastern and Southern Africa Centre of International Parasite Control (ESACIPAC), Kenya Medical Research Institute (KEMRI), Nairobi, Kenya; Neglected Tropical Diseases Support Center, Task Force for Global Health, Atlanta, GA, USA; London School of Hygiene & Tropical Medicine, London, UK; RTI International, Kampala, Uganda; Federal Ministry of Health, Addis Ababa, Ethiopia; Ministry of Health, Juba, South Sudan; The Carter Center, Juba, South Sudan; The Carter Center, Atlanta, GA, USA; Kamuzu University of Health Sciences, Lilongwe, Malawi; Amhara Regional Health Bureau, Bahir Dar, Ethiopia; Ministry of Health, Khartoum, Sudan; Sightsavers Kenya Country Office, Nairobi, Kenya; RTI International, Maputo, Mozambique; Task Force for Global Health, Addis Ababa, Ethiopia; Sightsavers, Haywards Heath, UK; Kirby Institute, UNSW Sydney, Sydney, Australia; University of British Columbia, Vancouver, Canada; Blantyre Institute for Community Outreach, Blantyre, Malawi; Muhimbili National Hospital, Dar es Salaam, Tanzania; Department of Ophthalmology, University of California San Francisco, San Francisco, CA, USA; World Health Organization Regional Office for Africa, Brazzaville, Republic of the Congo; Ministry of Health, Lusaka, Zambia; Frazer Institute, Faculty of Health, Medicine and Behavioural Sciences, The University of Queensland, Brisbane, QLD, Australia; The Carter Center, Khartoum, Sudan; University of Queensland Centre for Clinical Research, Brisbane, QLD, Australia; Ghana Health Service, Accra, Ghana; Ministry of Health, Lilongwe, Malawi; National NTD Control Program, Ministry of Health, Dodoma, Tanzania; Kongwa Trachoma Project, Kongwa, Tanzania; Neglected Tropical Diseases Program, Ministry of Health, Maputo, Mozambique; Department of Ophthalmology, University of Jos, Jos, Nigeria; Sightsavers, Nigeria Country Office, Abuja, Nigeria; University of Lusaka, Lusaka, Zambia; AFYAZEN Consulting, Nairobi, Kenya; Institute for Global Health Sciences, University of California San Francisco, San Francisco, CA, USA; Federal Ministry of Health and Social Welfare, Abuja, Nigeria; Ministry of Health, Kampala, Uganda; Global Health Division, International Development Group, RTI International, Durham, NC, USA; Department of Prevention, Control, and Elimination of Communicable Diseases, Pan American Health Organization, Washington, DC, USA; Ministério da Saúde, Maputo, Moçambique; Instituto de Higiene e Medicina Tropical, Lisboa, Portugal; Kenya National Public Health Institute (KNPHI), Division of Vector-Borne and Neglected Tropical Diseases Unit, Nairobi, Kenya; Wilmer Eye Institute, Johns Hopkins University, Baltimore, MD, USA; Noguchi Memorial Institute for Medical Research, University of Ghana, Accra, Ghana; Amhara Public Health Institute, Bahir Dar, Ethiopia; Department of Epidemiology and Biostatistics, University of California San Francisco, San Francisco, CA, USA; Malaria and Neglected Tropical Diseases Department, World Health Organization, Geneva, Switzerland

## Abstract

Trachoma is the leading infectious cause of blindness worldwide and is targeted for global elimination as a public health problem by 2030. As we approach this target, defining when population-level trachoma control interventions can be stopped, and determining how post-intervention surveillance should be performed, are becoming increasingly important. Molecular diagnosis of ocular *Chlamydia trachomatis* (*Ct*) using polymerase chain reaction (PCR) provides a highly sensitive and specific indicator of infection, yet there are currently no programmatic guidelines based on *Ct* infection. We assembled 86 cross-sectional surveys of ocular *Ct* infection from 13 countries, representing data from 46,914 children aged 1–5 years. Our analyses suggest that infection prevalence of 0.81% in this age group corresponds to a threshold below which population-level interventions can be stopped with at least 90% confidence. Correspondingly, with confidence exceeding 90%, we propose that interventions should continue when infection prevalence is greater than 1.99%. Interpretation of infection surveillance data is a generalisable challenge in low-prevalence settings for many infectious diseases. This study proposes a robust framework for how *Ct* infection surveillance could support accurate population-level assessments of trachoma endemicity and could allow a step-change in decision-making precision for programmes as we approach the endgame for trachoma elimination.

## INTRODUCTION

Trachoma, caused by ocular strains of *Chlamydia trachomatis* (*Ct*), is the leading infectious cause of blindness worldwide. As a neglected tropical disease (NTD), trachoma predominantly and disproportionately affects the world’s poorest communities and has been targeted for global elimination as a public health problem (EPHP) by 2030.^1^ Criteria for EPHP as specified by the World Health Organization (WHO) are threefold: a reduction in prevalence of trachomatous inflammation—follicular (TF; a clinical marker of conjunctival inflammation, associated with current or recent *Ct* infection) in children aged 1–9 years to less than 5% (TF_1-9y_ <5%) sustained for at least 2 years in the absence of ongoing MDA; reduction in prevalence of trachomatous trichiasis (TT; a painful sequelae of multiple trachoma infections that eventually leads to blindness) in ≥15-year-olds to less than 0.2%; and evidence that the health system can identify and manage incident TT cases. ^2^ Infection can be treated with a single dose of azithromycin, and antibiotic mass drug administration (MDA) has been central to co-ordinated and widely successful international efforts to eliminate trachoma over recent decades. As of July 2026, 32 of 78 formerly trachoma endemic countries have been validated by WHO for having achieved EPHP, with more expected to do so over the coming years.^3–6^

The conjunctival scarring which instigates the pathological processes that lead to TT and trachomatous blindness only becomes clinically significant after multiple ocular *Ct* infections over many years^7^. The implication of this is that there will be a point at which *Ct* transmission intensity is so low that population-wide interventions such as MDA are not necessary to prevent blindness. Furthermore, to date, in countries where EPHP has been validated (i.e. TF_1-9y_ has been sustained at <5% for a minimum of two years), resurgence of TF has not been reported. This suggests that whilst TF is not a direct marker of infection, the 5% threshold may correspond to, or even be more stringent than, an infection prevalence breakpoint, defined as a level of *Ct* infection below which transmission is not sustained.

As we approach the final stages of eliminating trachoma as a public health problem globally, the so-called elimination endgame, there is increasing interest in the use of surveys which include sampling and assays for ocular *Ct* infection to evaluate the epidemiology of trachoma at district level, and ultimately to support decision-making for national elimination programmes.^8–13^ Detection of *Ct* DNA on conjunctival swabs using polymerase chain reaction (PCR) testing provides a highly sensitive and specific indicator of current infection.^14^ This is in contrast to TF, an imperfect proxy for *Ct* infection, which can persist for months after infection is cleared. There are also justifiable concerns regarding the specificity and sensitivity of TF diagnosis, and the risk that even well-trained examiners may become less consistent at very low prevalence levels (a phenomenon known as grader drift), potentially increasing the rate of misclassification errors.^15–17^

Despite the growing availability of established, commercially-available PCR platforms for detection of *Ct* infection, key gaps remain regarding how to include estimation of ocular *Ct* infection prevalence in population-based trachoma surveys, and how results should be interpreted, in particular with regards to the decision to stop population-wide MDA.

We hypothesise that as formerly-endemic populations approach the EPHP thresholds, the prevalence of *Ct* infection will approach zero in children aged 1–5 years. This age group is considered the core transmission group for trachoma, and episodes of infection are generally longer in this age group compared to older individuals.^7,18–21^ Prevalence of *Ct* infection in 1–5-year-olds should therefore give a representative picture of transmission dynamics sufficient to support accurate population-level assessments of trachoma endemicity.

Here, we assembled 86 cross-sectional surveys of ocular *Ct* infection across a gradient of endemicities in 13 countries, together with contemporaneous population-level measures of trachoma clinical signs and serological markers, representing data from 60,559 children aged 1–9 years (46,914 aged 1–5 years) collected between 2007 and 2024. Our primary objective was to use these collated datasets to characterise *Ct* infection epidemiology that corresponds to a need for population-level action with a sufficient level of confidence to support decision-making.

Applying Bayesian modelling approaches to the assembled empirical datasets, we have identified candidate infection prevalence thresholds for implementing and for stopping population-level trachoma control activities, and have used a mechanistic model-based simulation approach to explore the influence of survey timing in relation to previous rounds of MDA.

Interpretation of infection surveillance data is a critical and generalisable challenge in low-prevalence and near-elimination settings for many infectious diseases. Our approach is broadly applicable to other diseases for which interventions are delivered at population level, in particular other NTDs for which preventative chemotherapy is the core strategic intervention.^22^ Our results provide new, data-driven guidance for use of PCR and *Ct* infection prevalence in trachoma surveillance as we approach global elimination of trachoma as a public health problem.

## RESULTS

### Characterising trachoma infection prevalence distributions corresponding to need for population-level interventions

Building on a recently published methodological framework characterising seroconversion rates (SCR) which correspond to the need for intervention against active trachoma,^23^ our approach in identifying thresholds for programmatic action was analogous to using prevalence of *Ct* ocular infection in children aged 1–5 years as a population-level diagnostic tool. When characterising cut-offs levels for diagnostic tools such as biomarkers, reference groups of diseased and non-diseased individuals are typically defined based on available diagnostic tests and presence/absence of clinical signs.^24^ In a comparable way, we aimed to identify populations within our assembled datasets that could be used as reference populations-those with a high level of transmission corresponding to a very clear need for trachoma-specific interventions such as MDA in order to prevent future blindness, and those where such interventions are clearly not needed.

Of the 86 population-level surveys that met the inclusion criteria (Supplementary Figure S1), 30 were identified as having a high probability that interventions would be needed to prevent blindness, categorised as “Action needed”, and 31 were identified as having a high probability that population-level interventions would not be needed to prevent blindness, categorised as “No action needed” (Table 1). These categorisations were based on consensus opinion of 41 expert reviewers (*Methods*) and based on *Ct* infection prevalence together with all available contemporaneous clinical and serological data for each population (*Methods*). The remaining 25 surveys were categorised as “Unclassified”.

**Table 1.** Summary of *Chlamydia trachomatis* infection prevalence, trachomatous inflammation—follicular (TF) prevalence, trachomatous inflammation—intense (TI) prevalence, seroprevalence of Pgp3 IgG, seroconversion rate (SCR) by survey and survey categorisation. (See appended spreadsheet) CrI= Bayesian Credible Intervals Table 1 link: https://doi.org/10.6084/m9.figshare.34892772

Each survey population was treated as an evaluation unit (EU), which is generally an administrative unit representing 100,000–250,000 individuals and is the WHO-recommended population unit in which trachoma elimination activities take place. ^25^

The included datasets spanned a broad range of *Ct* infection prevalence levels (median of EU-level prevalence ranged from 0.03% to 36%, Figure 1). When prevalence data for EUs within each category were combined to generate pooled prevalence density distributions (Figure 2), there was a region of overlap in the 0–2% prevalence range. The two distributions are however clearly distinct, with the majority of the “No action needed” distribution falling below 1% prevalence, a much wider prevalence distribution for the “Action needed” category, and a separation between the two categories for the majority of the prevalence range.

**Figure 1.**
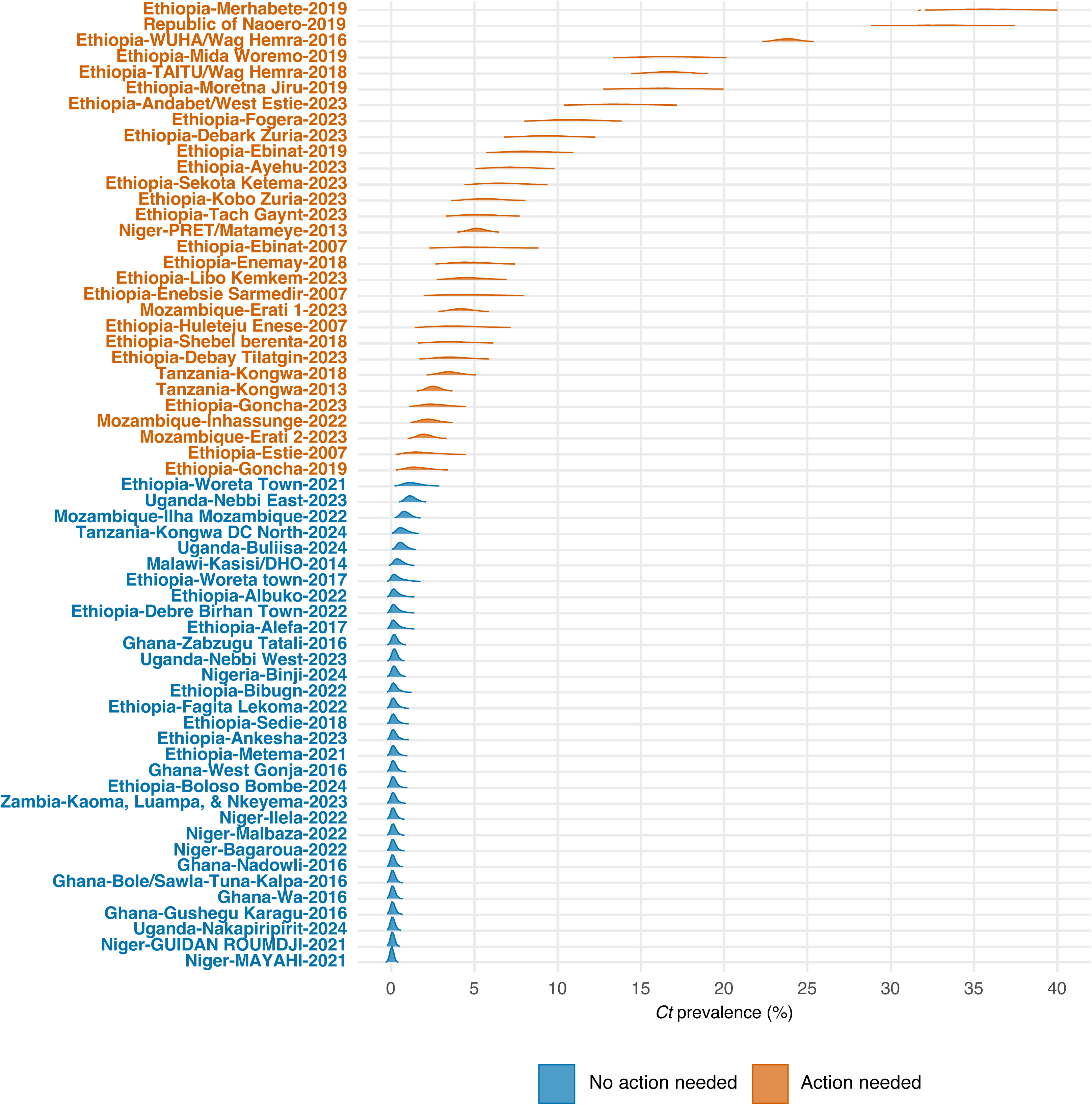
*Ct* infection prevalence (%) density distributions. Density distributions for 61 classified evaluation units, with colour representing category “Action needed” (orange) and “No action needed” (blue) respectively. N=45,240. Corresponding plots including unclassified evaluation units are given in Supplementary Figure S2.

**Figure 2.**
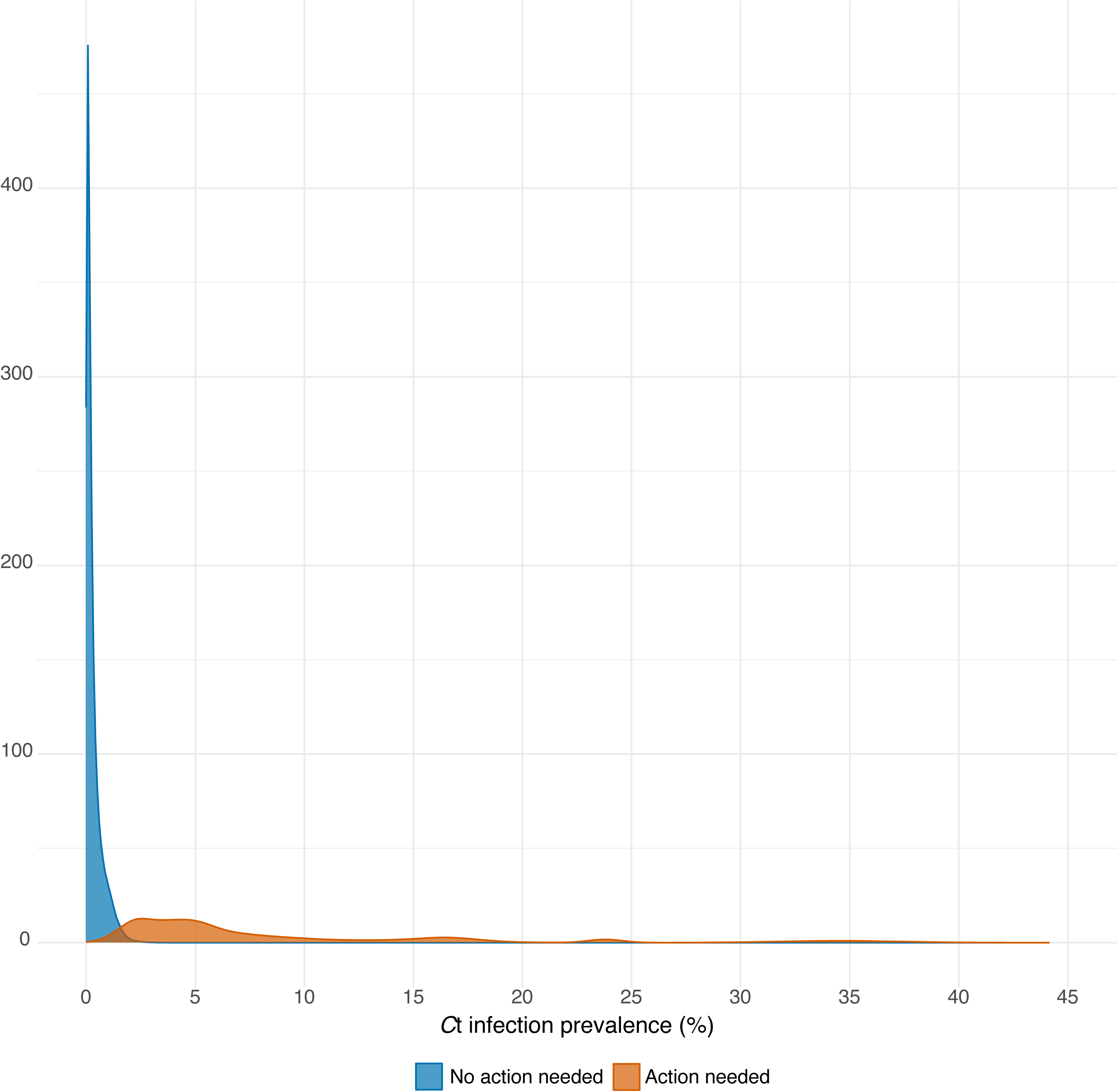
Pooled probability density distributions for *Ct* infection prevalence (%) by category “Action needed” (orange) and “No action needed” (blue).

### Probability of programmatic category as a function of infection prevalence and identifying prevalence thresholds corresponding to programmatic decisions

Our primary objective was to use the empirical datasets to identify regions of high probability of the need for population-level programmatic action (or not) based on *Ct* infection prevalence, and from those results develop candidate thresholds for decision-making. Following the Bayesian mixture model methods of Kamau *et al*., 2025,^23^ we multiplied the pooled probability distributions for each category (Figure 2) with a prior probability to generate a posterior probability for each category as a function of prevalence (*Methods*).

Using uninformative priors (i.e. assuming equal prior probability of each programmatic category), we derived posterior probabilities for category as a function of *Ct* infection prevalence in children aged 1–5 years (Figure 3). The probability that no programmatic action is required exceeded 90% when the EU-level *Ct* prevalence was ≤0.81%, and exceeded 95% when prevalence was ≤0.63%. Conversely, the probability that transmission is at a level that requires population-level interventions exceeded 90% when infection prevalence was ≥1.99% and exceeded 95% when prevalence was ≥2.19%.

**Figure 3.**
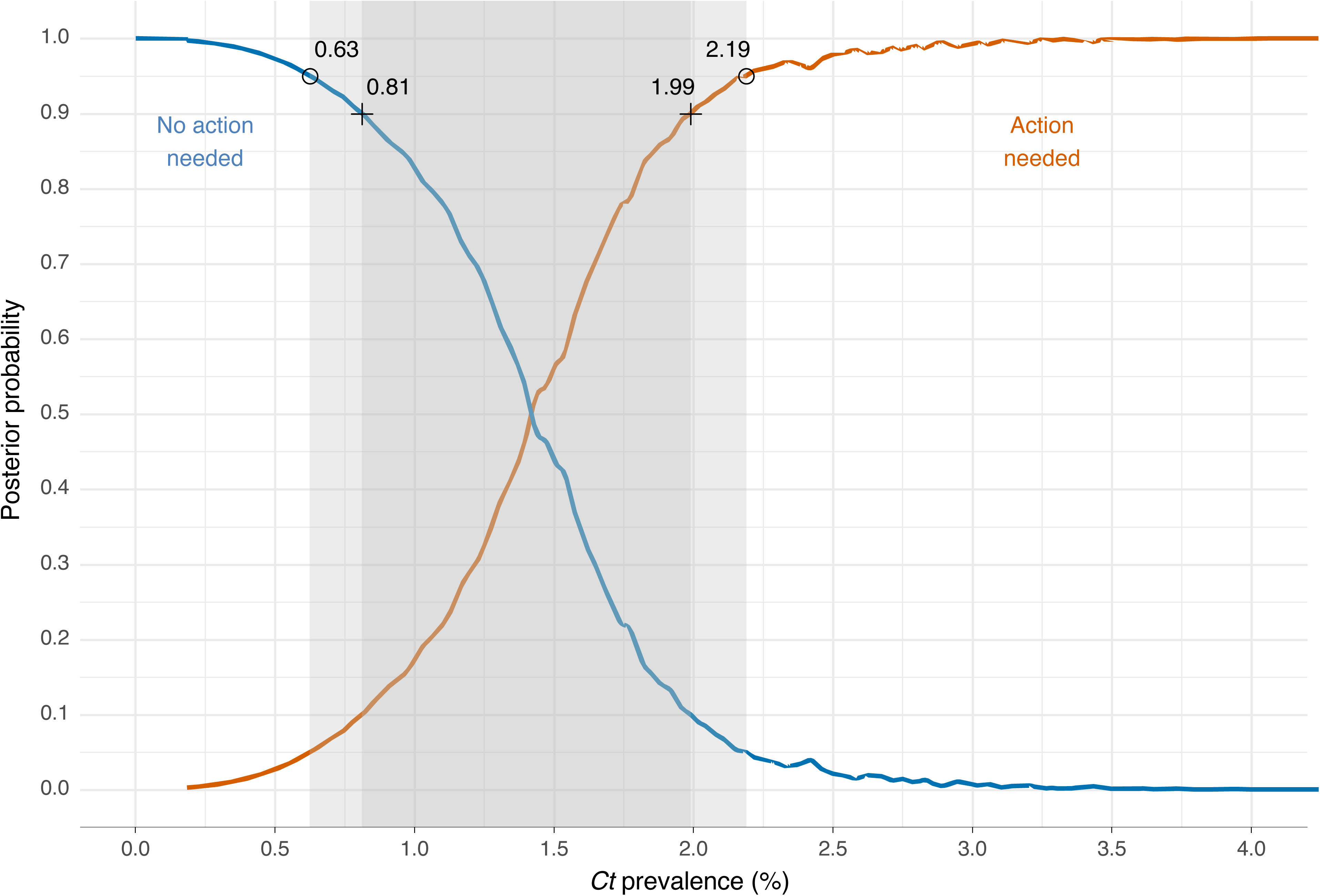
Posterior probabilities of programmatic category “No action needed” (blue) or “Action needed” (orange) given prevalence of *Ct* infection in children aged 1-5 years. Uninformative priors have been assumed (equal prior probability for each category). 90% (cross) and 95% (circle) probability thresholds are highlighted, with shaded area representing the range of prevalence values with less than 90% (dark grey) or 90-95% (light grey) probability of either category. Corresponding thresholds for informative prior scenarios are given in Table 2 and posterior probability plots for informative prior scenarios are given in Supplementary Figures S3 and S4.

**Table 2.**
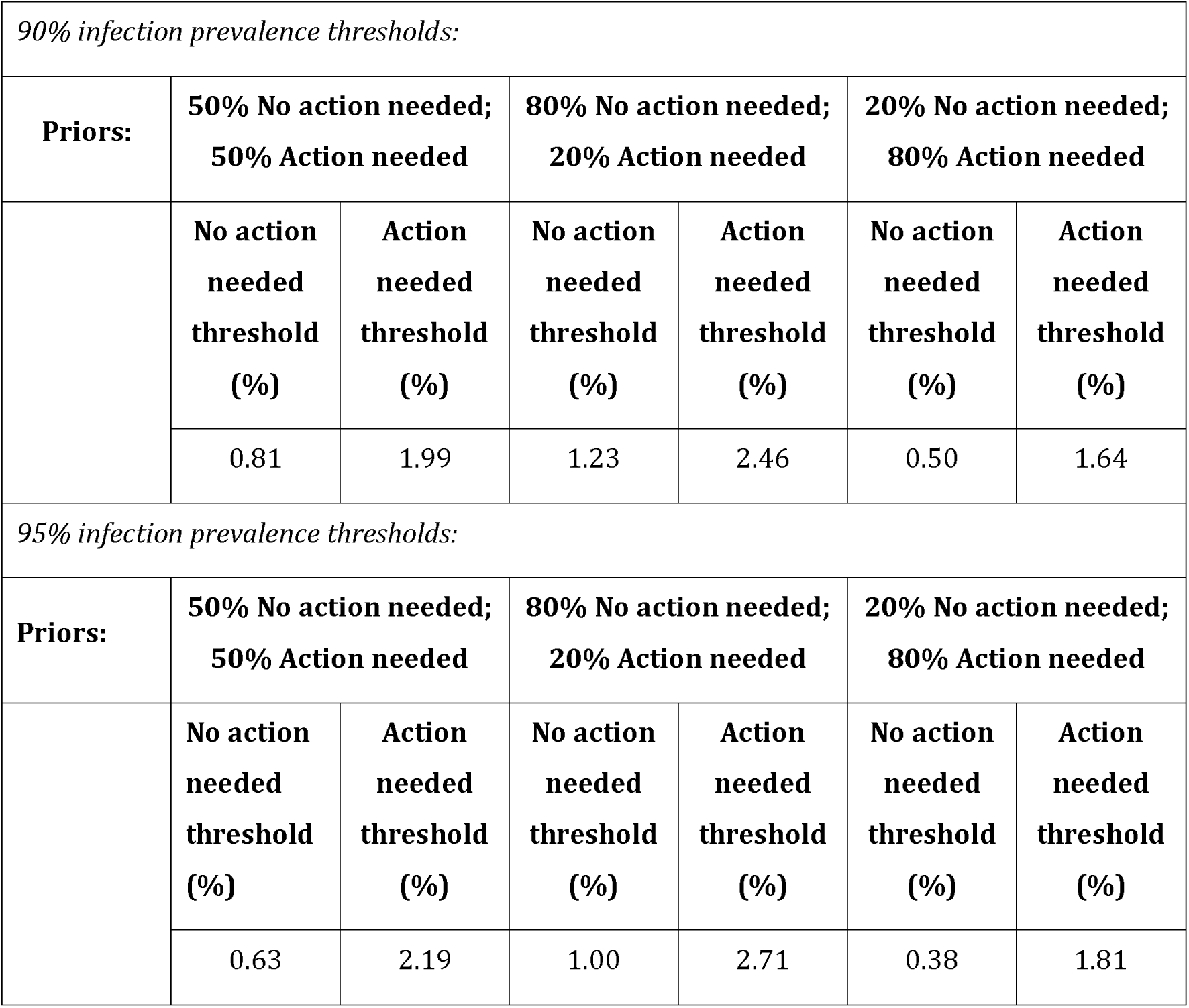
Summary of *Ct* prevalence thresholds that correspond to 90% or 95% probability of category, “Action needed” or “No action needed”, with given prior probabilities. Below the lower “No action needed” infection prevalence threshold, the probability of an evaluation-unit (EU) not needing population-level interventions exceeds 90% (upper panel) or 95% (lower panel). Above the upper “Action needed” infection prevalence threshold the probability of an EU needing population-level interventions (such as mass-drug administration) to prevent blindness exceeds 90% (upper panel) or 95% (lower panel). The 50/50 prior corresponds to an uninformative prior probability, and the 80/20 priors correspond to informative prior probabilities of 80% “No action needed” vs 20% “Action needed” and vice versa.

| *90% infection prevalence thresholds:* |  |  |  |  |  |  |
|---|---|---|---|---|---|---|
| **Priors:** | **50% No action needed; 50% Action needed** |  | **80% No action needed; 20% Action needed** |  | **20% No action needed; 80% Action needed** |  |
|  | **No action needed threshold (%)** | **Action needed threshold (%)** | **No action needed threshold (%)** | **Action needed threshold (%)** | **No action needed threshold (%)** | **Action needed threshold (%)** |
|  | 0.81 | 1.99 | 1.23 | 2.46 | 0.50 | 1.64 |
| *95% infection prevalence thresholds:* |  |  |  |  |  |  |
| **Priors:** | **50% No action needed; 50% Action needed** |  | **80% No action needed; 20% Action needed** |  | **20% No action needed; 80% Action needed** |  |
|  | **No action needed threshold (%)** | **Action needed threshold (%)** | **No action needed threshold (%)** | **Action needed threshold (%)** | **No action needed threshold (%)** | **Action needed threshold (%)** |
|  | 0.63 | 2.19 | 1.00 | 2.71 | 0.38 | 1.81 |

These thresholds define three programmatically relevant regions: a low prevalence range in which the cessation of MDA can be supported with at least 90% confidence, a higher prevalence range in which continued intervention is indicated with at least 90% confidence, and an intermediate range in which there is less than 90% certainty of either category (highlighted as dark grey in Figure 3).

We considered the thresholds generated from this uninformative prior scenario to be most applicable to populations for which there are no prior data to inform decision-making or for which such data are equivocal. We also considered scenarios in which prior information strongly indicated either ongoing transmission or transmission control, using priors of 80% “Action needed” *vs*. 20% “No action needed”, and vice versa, respectively (*Methods*). Prior information could, for example, include prevalence of anti-*Ct* antibodies, prevalence of TF, or presence of end-stage trachoma outcomes such as TT, particularly in an EU that has never received MDA.

With a prior probability of “Action needed” given as 80%, a prevalence threshold of ≤0.5% was required to exceed 90% probability of no programmatic action (Table 2). This reflects a stricter criterion for deciding not to intervene in settings for which other indicators are suggestive of an ongoing need for interventions. Thresholds identified for all priors are summarised in Table 2, with corresponding posterior probability plots for the informative prior scenarios shown in the Supplementary Materials (Supplementary Figures S3 and S4).

### Impact ofsurvey timing post-MDA: Sensitivity analyses

Over the course of a programme of MDA, which typically comprises one to five annual rounds of population-wide treatment, there is expected to be a significant drop in infection prevalence, which can ultimately lead to interruption of transmission. Interruption may be preceded by a period of local sub-critical transmission, leading to stuttering chains (self-limiting local chains of transmission that fade to extinction) or a period of low-level transmission.^26^ Alternatively, after MDA there may be an initial decrease in prevalence, but ongoing transmission ultimately leads to a resurgence in prevalence to a level at which MDA is necessary. Survey timing may therefore influence how we should interpret infection prevalence estimates after MDA: if an infection prevalence survey is carried out too soon, it may not yet be clear whether transmission is fading towards interruption or if there is ongoing transmission warranting further MDA rounds.

Of the 86 datasets, 84 were from surveys carried out in populations that had a history of receiving MDA and two were baseline surveys from populations with no history of prior MDA (Republic of Naoero 2019 and South Sudan, Torit, 2024). Of the 84 surveys in which there was a history of MDA, four were from randomised controlled trials where timing and number of rounds of MDA varied within the dataset by study arm. For the remaining 80 populations with history of EU-wide MDA, the median time from MDA to survey was 10 months (ranging 7 months to 13 years).

We re-calculated the thresholds corresponding to 90% probability of “Action needed” or “No action needed” for subsets of survey data according to the timing of MDA relative to the survey, using uninformative priors. For populations with no history of MDA or in which MDA was carried out ≥12 months prior to the survey (n=36), the *Ct* prevalence threshold corresponding to 90% probability of “No action needed” was estimated as 1.08% in children aged 1–5 years. This was higher than the corresponding 90% threshold for surveys carried out <12 months after the last MDA (n=46), estimated to be 0.80%.

Results of full sensitivity analyses according to survey timing relative to MDA are given in Supplementary Table S1.

### Impact ofsurvey timing post-MDA: Modelling approach

Decisions on whether to halt or continue population-level interventions after a programme of MDA are currently based on TF_1-9y_ at the time of the trachoma impact survey (TIS). It is recommended that the TIS be carried out between 6 and 12 months after the last planned MDA. If the criteria for EPHP are met (TF_1-9y_ <5%), no further MDA or surveillance is generally carried out until the trachoma surveillance survey (TSS) at least two years later.^27^ If infection prevalence is to be used to inform decisions during the 6–12-month TIS window, it is therefore critical that slowly resurging infection in the population does not lead to misclassification as “No action needed”. We used model simulations fitted to the two categories of empirical prevalence distributions (“Action needed” and “No action needed”) at 12 months post-simulated MDA and generated the posterior probabilities for programmatic category as a function of prevalence at weekly timesteps between 6 and 13 months post-MDA (*Methods*). The density plots for the two programmatic categories generated from model-simulated data show a greater degree of overlap at 6 months post-MDA as compared to 12 months post-MDA (Figure 4). From time-specified posteriors, we calculated time-adjusted prevalence thresholds corresponding to ≥90% and ≥95% probabilities for each programmatic category at each timepoint. We used locally estimated scatterplot smoothing (LOESS) to fit a regression line through these estimates to generate time-adjusted threshold curves representing 90% and 95% thresholds for “No action needed”. The 90% and 95% thresholds increase consistently from 0.35% and 0.11%, respectively at 6 months post-MDA (Figure 5), converging with the corresponding 90% and 95% empirical threshold estimates of 0.81% and 0.63% at approximately 10 months post-MDA. Example time-adjusted point estimates of programmatic thresholds used to generate the 90% curve are given in Supplementary Figure S7.

**Figure 4.**
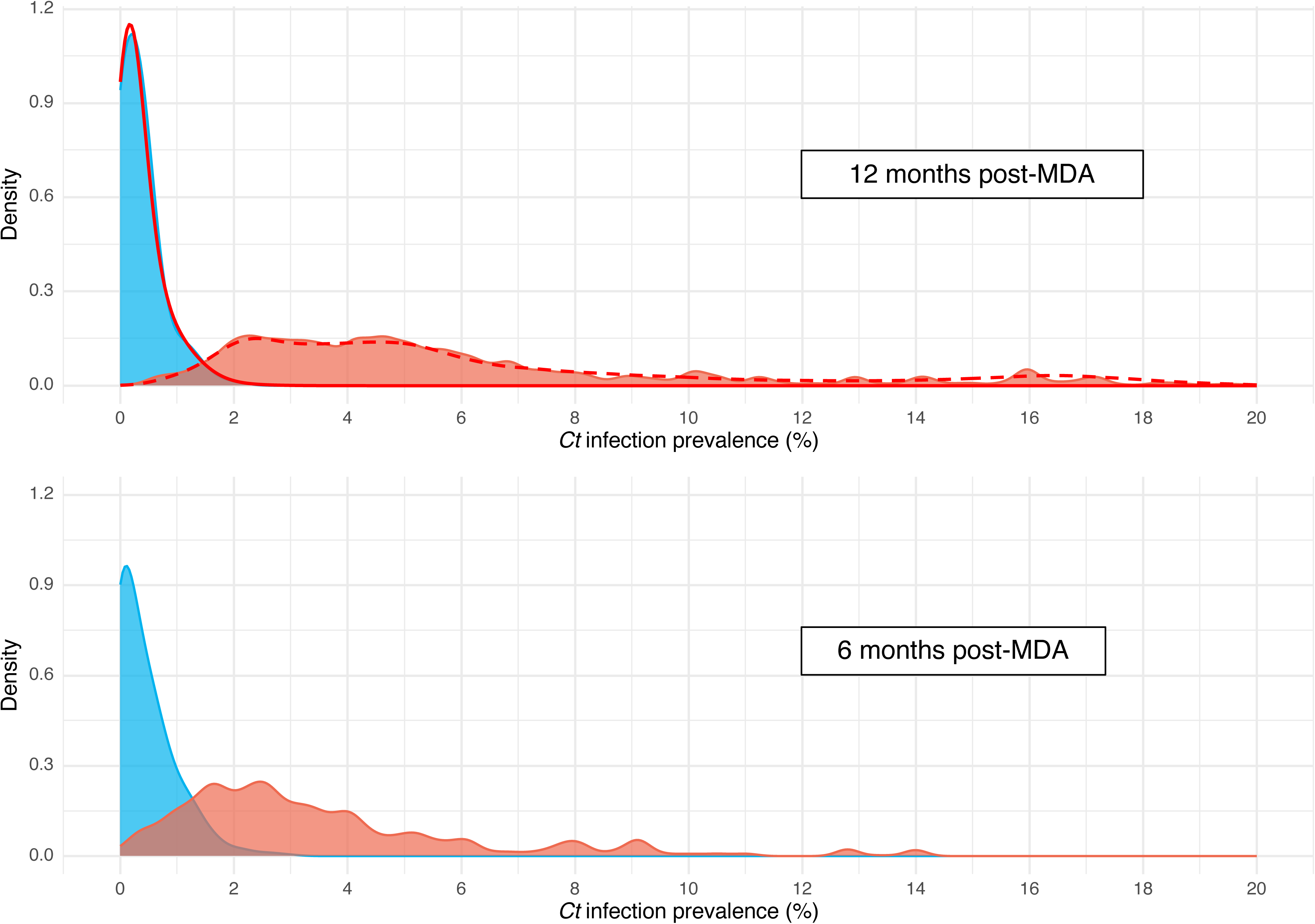
Pooled density distributions of Ct infection prevalence in children aged 1-5 years (%) for simulated datasets at 12 months and 6 months post-MDA. Modelled scenario categories representing “Action needed” (orange) and “No action needed” (blue). Empirical density distributions used to fit the model at 12 months shown as red outlines (dashed=Action needed, solid=No action needed).

**Figure 5.**
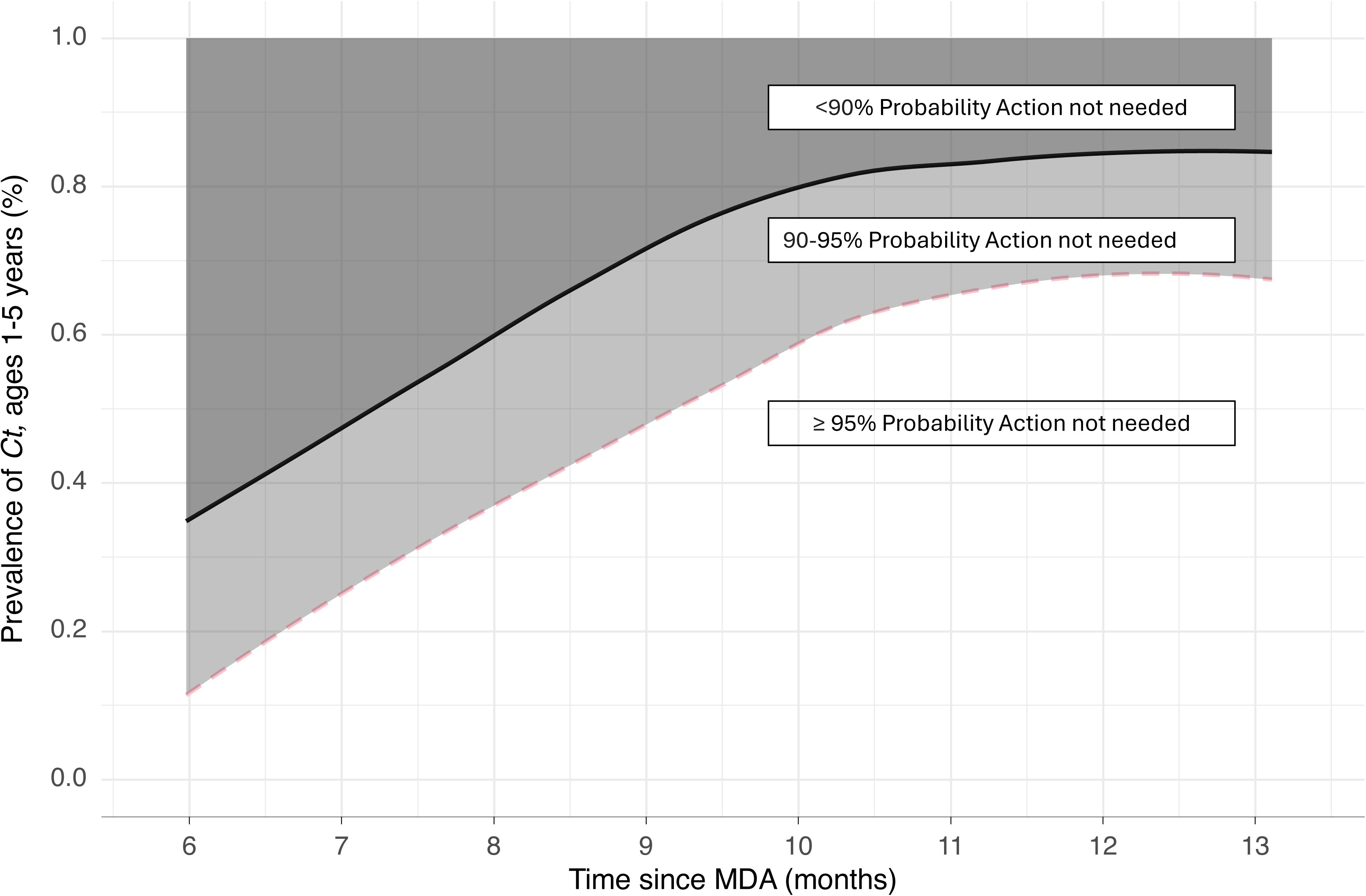
Time-adjusted Ct infection prevalence thresholds for category “No action needed” after a programme of mass drug administration (MDA). The black line represents the 90% threshold and the pink dashed line represents 95% threshold. The dark grey region therefore corresponds to less than a 90% probability of no programmatic action needed, the light grey area corresponds to a 90–95% probability of no action needed, and the white area 95% or greater probability of no action needed. Thresholds were estimated using uninformative priors. Corresponding plots including 85% and 85% thresholds are given in Supplementary Figure S9.

The corresponding time-adjusted thresholds for ≥90% probability of category “Action needed” showed little change between 6 and 12 months. Furthermore, it would not be necessary to time-adjust the “Action needed” threshold if a more conservative interpretation of the thresholds is applied, in which all EUs with infection prevalence above the “No action needed” threshold are considered for treatment.

### Probability of Type 1 and Type 2 error: “Under-treatment” and “Over-treatment”

We consider that there are two categories of error when making decisions on whether to halt or continue with community-level interventions after a programme of MDA. If the default position is to continue with MDA, a Type 1 error would correspond to “Under-treatment”, i.e. incorrectly stopping MDA when further intervention is needed, and a Type 2 error would correspond to “Over-treatment”, i.e. continuing MDA when it is not needed.

We compared the median *Ct* infection prevalence for all classified EUs to the proposed 90% probability thresholds of 0.81% for no programmatic action, and 1.99% for programmatic action needed. For EUs classified “Action needed”, 93% (28/30) had a median prevalence estimate greater than 1.99%, and 100% had a median prevalence estimate greater than 0.81%. Of EUs classified “No action needed”, 90% (28/31) of median prevalence estimates were below the 90% probability threshold, when using the time-adjusted threshold (Figure 6) and the non-time adjusted threshold of 0.81%. No EUs categorised as “No action needed” had an infection prevalence above the 1.99% threshold.

**Figure 6.**
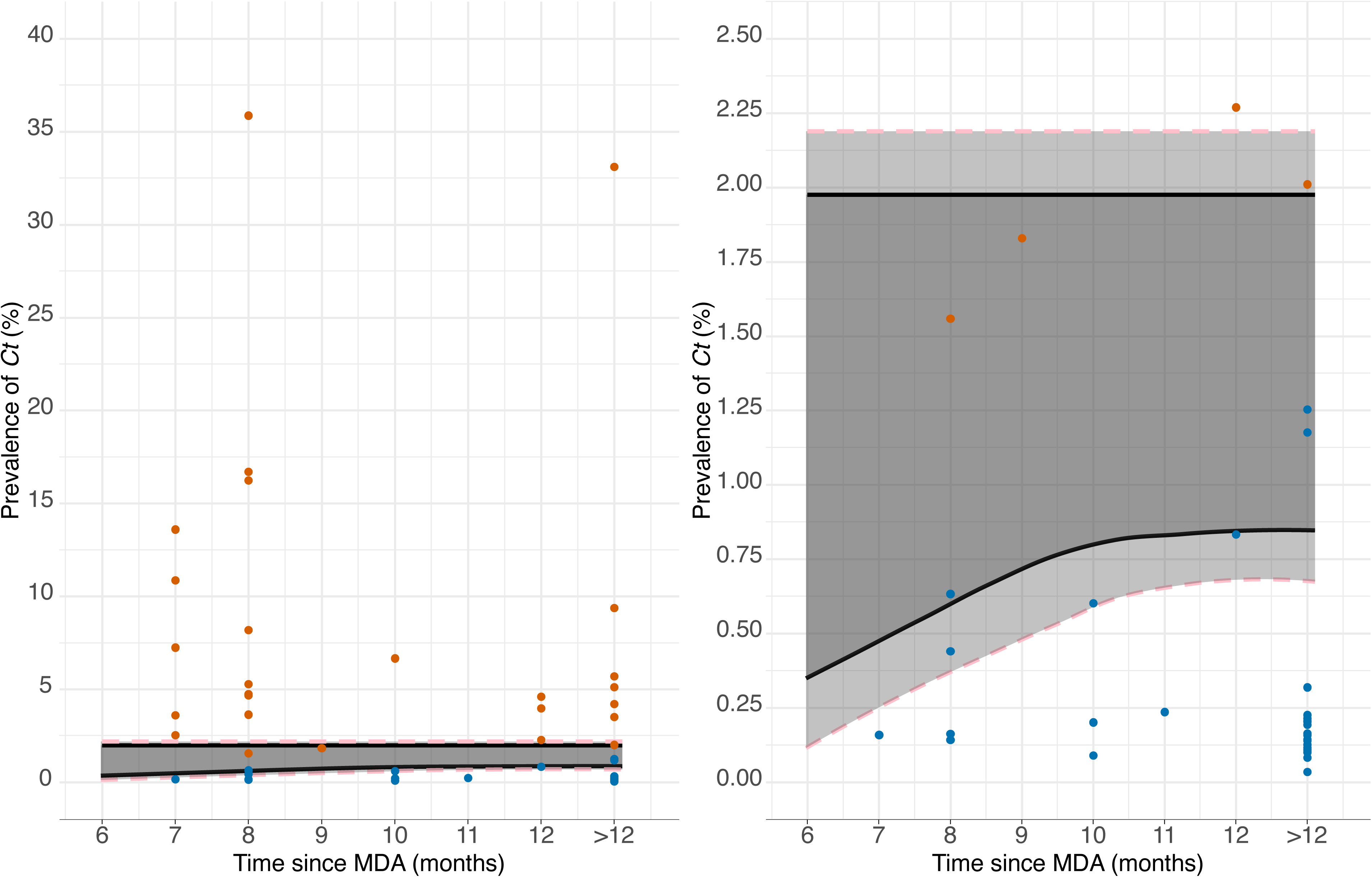
Prevalence point estimates for Ct in children aged 1–5 years compared to time-adjusted thresholds. In panel A, the median of prevalence posteriors for all classified EUs (N=61) is plotted by time post-MDA compared to 90% programmatic action thresholds (black line) and 95% thresholds (pink dashed line). Point prevalence estimates for EUs classified as “Action needed” are shown as orange circles, and for EUs classified as “No action needed” shown as blue circles. The dark grey region corresponds to less than a 90% probability of either category, the light grey area corresponds to a 90–95% probability of “Action needed” (upper light grey region) or “No action needed” (lower light grey region). The range of intermediate prevalence values where there is less than a 90% probability of either programmatic category is relatively narrow, with the majority of prevalence values falling within the >90% probability range. Prevalence for five out of 61 EUs (8%) falls within the intermediate range, none of the “Action needed” EUs have a prevalence estimate less than the proposed thresholds for no programmatic action, and none of the “No action needed” EUs have a prevalence estimate greater than the threshold for programmatic action. Decisions on whether to continue interventions when prevalence falls within the <90% zone after MDA will therefore be a pragmatic trade-off between risk of over-treatment and under-treatment. Panel B represents the same data as panel A but focusing on the 0–2.5% region.

The proposed thresholds for programmatic categories allow for an intermediate range in which there is a less than 90% probability for either category. Prematurely halting MDA (under-treatment) has the potential for a significant negative outcome, i.e., unchecked resurgence of transmission. If we therefore apply a more conservative interpretation of the thresholds to the classified data, in which MDA is continued if prevalence is more than the “No action needed” threshold (even if it is below the “Action needed” threshold), there would be zero cases of under-treatment using the classified datasets and a 10% probability of over-treatment (three “No action needed” EUs were between 0.81% and 1.99%).

### Probability of programmatic category for unclassified evaluation units

A potential use-case scenario for infection prevalence thresholds is when measurements of other indicators, specifically TF_1–9y_ and SCR, are discordant or there is unusual trachoma epidemiology in a population. To demonstrate this application of infection prevalence thresholds to determine programmatic action, we used the 25 “unclassified” EUs (those that did not have a consensus expert opinion on programmatic action category). For each EU, we compared the median of posterior prevalence distribution to the candidate programmatic action thresholds (Figure 7A). Using the 90% uninformative scenario thresholds of 1.99% and 0.81% for “Action needed” and “No action needed” respectively, two unclassified EUs could be re-classified as “Action needed” (Mozambique-Mossuril-2022 and Mozambique-Nacala-A-Velha-2022) and 14 EUs could be re-classified as “No action needed”.

**Figure 7.**
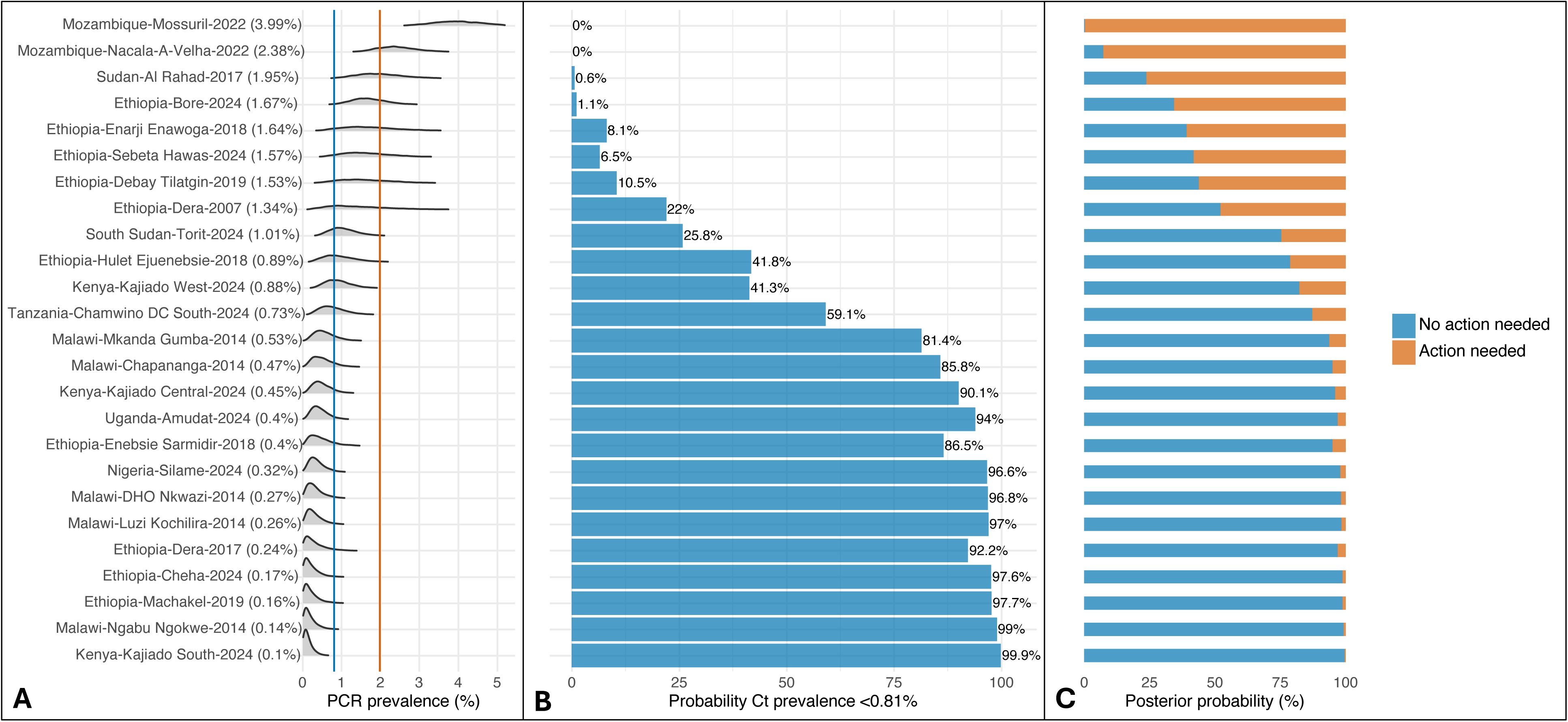
Probabilistic estimation of need for programmatic action for unclassified EUs. 25 EUs were designated “Unclassified” due to not reaching the thresholds for consensus in the expert survey. The posterior prevalence distributions for unclassified EUs are plotted with the uninformative 90% “Action not needed” and “Action needed” thresholds of 0.81% (blue) and 1.99% (orange) respectively (Panel A). The proportion of these prevalence distributions that fall below the “No action needed” threshold of 0. 81% is given in Panel B. The posterior probability of each programmatic category is given in Panel C.

The remaining nine EUs fell between the two thresholds considered here. We note that only one of these was from a survey carried out ≥12 months after MDA (Supplementary Figure S8).

We considered two approaches to informing categorisation of EUs as alternatives to comparing point estimates to a single threshold. These may be applicable when point estimates fall between the two programmatic thresholds, when other data indicate disparate categorisations, or when additional confidence is required, for example, the final surveillance survey before declaring EPHP. First, we calculated the probability that the prevalence for each “Unclassified” EU was below the 90% “No action needed” threshold of 0.81% (Figure 7B), and secondly, we used the pooled distributions of the categorised EUs combined with the posterior prevalence distribution for each unclassified EU to generate posterior probabilities of programmatic category (Figure 7C). The summary results from the expert opinion survey together with posterior probabilities generated here are given in Supplementary Table 2.

### Further sensitivity analyses

We considered the potential for bias by running jack knife n-1 sensitivity analyses, excluding i) each country in turn and ii) each EU in turn, and re-calculating the posterior probabilities of “Action needed” and “No action needed”. These analyses indicated that Ethiopia was highly influential in the analyses but that there was no bias due to any single EU-level dataset (Supplementary Figures S10 and S11). We carried out further sensitivity analysis to assess the impact of including children aged 6–9 years in the analyses (Supplementary Figures S12 and S13). These results indicate that there is no need to age-adjust *Ct* prevalence estimates when surveys include only children aged 1–5 years.

## DISCUSSION

Our results demonstrate how prevalence of *Ct* infection in children aged 1–5 years could inform decision-making for trachoma programmes as we approach the elimination endgame. Building on the framework proposed by Kamau *et al* 2025^23^ and using an extensive dataset comprising 86 ocular *Ct* infection surveys from 13 countries and a wide range of populations, we present a quantitative framework for interpreting *Ct* prevalence in children aged 1–5 years in the context of trachoma elimination. We have identified infection prevalence thresholds corresponding to high probabilities that population-level intervention was either required or not required, demonstrated how these may be adjusted both in the context of other data and the timing of the survey after MDA, and shown how probabilistic approaches can support decision-making when prevalence estimates fall close to decision boundaries. Together, our findings provide an evidence-based framework for incorporating PCR-based infection surveys into decision-making for trachoma elimination programmes.

For trachoma, as for many infectious diseases, misclassification error can become a greater challenge for decision-making in low-prevalence or near-elimination settings. TF_1-9y_ is currently the primary metric for trachoma elimination programmes, determining where MDA should be implemented, informing Pfizer’s azithromycin donation programme, defining when population-level treatment can be stopped, and is used for assessing whether EPHP has been achieved. TF is, however, an indirect measure of current or recent ocular *Ct* infection and has well-documented limitations regarding its sensitivity and specificity, and there are concerns regarding the risk of grader drift at low prevalence levels.^15,16,28^ In contrast, diagnostic assay-based approaches, including PCR detection of *Ct* DNA on conjunctival swabs and serological detection of IgG response to the *Ct* antigen Pgp3, are highly sensitive and specific regardless of prevalence. In recent years, PCR assays for *Ct* have become far more widely available, as has interest in their use in trachoma surveys. Part of this interest has stemmed from growing awareness of the phenomena of persistent and recrudescent TF, and the potential contribution to these phenomena of the time-lag between TF and clearance of infection, as well as inconsistent clinical grading, despite intensive international focus on standardization and certification of graders.^29,30^

Children aged 1–5 years are the core transmission group for trachoma,^18,31^ and prevalence of infection in this age group should provide an informative representation of current transmission. We estimated thresholds for programmatic categories corresponding to 90% and 95% probability, recognising that acceptable levels of confidence may vary by context. It is important to acknowledge that risk is a function of both probability and impact, and the potential impact of incorrect decisions includes delayed/missed MDA (“under-treatment”), MDA when it is not necessary (“over-treatment”), or missed resurgence.

Our analyses of longitudinal village-level post-MDA infection prevalence data from Niger and Ethiopia^19,32–37^ (Supplementary Materials) indicated that when applying our proposed thresholds, probability of under-treatment was higher at 6 months post-MDA (15%) than at 12 months post-MDA (<4%). Given that it is recommended for trachoma impact surveys to be carried out 6–12 months after the last planned MDA, these findings imply a pragmatic trade-off between reducing probability of misclassification and ensuring timely intervention. Delaying a survey increases the likelihood that post-MDA resurgence will be apparent, but also risks delaying further MDA where it is needed. We applied a transmission modelling approach to propose time-adjusted thresholds for the 6–12-month TIS window. Modelling results indicated that by approximately 10 months post-MDA, the non-time-adjusted 90% threshold of 0.81% for “No action needed” should be applicable. If a threshold of 0.63% is applied, modelling output indicated that by 8 months this would correspond to at least a 90% probability of category “No action needed”, and by approximately 10 months this would correspond to at least 95% probability of the “No action needed” category. The overall implication of these results is that if surveys are carried out before 10 months post-MDA, a lower prevalence threshold may be needed to have the same confidence in a “No action needed” decision, or alternatively, if a fixed decision-making threshold is defined, confidence around this threshold may need to be adjusted.

Our aim here was not to define interruption of *Ct* transmission, but the modelling approach described could be adapted to identify potential breakpoints (prevalence levels below which transmission is likely to decline to zero) for given time-points post-MDA. Outside of clinical trials, trachoma programmes do not typically re-sample populations between TIS and TSS, but future work could include repeat sampling of communities to better understand the dynamics of sub-critical trachoma transmission and resurgence. The resulting data could validate our proposed time-adjusted thresholds.

Our approach delineates an intermediate prevalence range in which there is less than 90% probability of either programmatic category (“Action needed” or “No action needed”; 0.81–1.99% under the uninformative prior scenario). When prevalence estimates fall within this range, it may be appropriate to repeat surveys, examine additional infection indicators such as anti-Pgp3 antibodies, or consider contextual information such as timing since MDA. However, where a binary decision is required, a conservative strategy in which population-level interventions are continued when prevalence falls within the intermediate range, would lead to low levels of under-treatment, with a corresponding trade-off of increased probability of over-treatment (estimated 10% overtreatment if populations in the intermediate range are treated vs. 7% under-treatment if populations in the intermediate range are not treated). We also demonstrate two probabilistic approaches that may aid interpretation of surveys in the intermediate range: estimating the probability that prevalence is below a given threshold, and estimating the posterior probability of each programmatic category based on the full prevalence distribution, rather than the point estimate alone.

These approaches could also provide context for difficult-to-interpret data. For example, if the distribution of prevalence in an EU-level survey is highly heterogenous with positive cases clustered within a single village, this would be seen as a positively skewed prevalence distribution with a low point prevalence estimate. In such cases, quantifying the probability of the need for programmatic action may be more informative than simple comparison of point prevalence estimates to given thresholds. This approach is highly applicable to decision-making for other diseases, in particular in resource-poor settings, where cost of surveillance can become a disproportionate cost to programmes, and repeat surveys if results are equivocal may not be feasible.

The bounds of the intermediate range we have defined as having less than 90% probability of either category are relatively narrow, which raises the question of whether misclassification error at individual level could lead to misclassification at population level. Published laboratory validation studies of commercially available PCR platforms report very high sensitivities and specificities in the range of >98–100%,^14^ but field conditions may be expected to reduce test performance through delays in sample transport, cold-chain issues, contamination, or other operational factors. Importantly, any real-world variability in test performance is inherently incorporated into our analyses. Our proposed thresholds are derived from empirical surveys undertaken in field conditions, and as such should be considered applicable to field-observed data. The surveys used in our analyses were mainly carried out as part of routine programme implementation work, with some carried out primarily in a research context (for example, cluster randomised trials). In both cases, the data originated from surveys with strong quality assurance and quality control measures, using commercially-available externally-validated PCR platforms. If outputs from future surveys are to be interpreted against the thresholds proposed here, use of equivalent PCR platforms, together with standardisation of sample collection, laboratory processing, and prevalence estimation will be crucial. ^30^ The datasets analysed here are not a random subset of all trachoma-endemic populations. For example, some surveys carried out more than 2.5 years after MDA were surveillance surveys, indicating that TF_1-9y_ was already less than 5% at an earlier TIS, and in some other cases surveys incorporating swab collection for PCR may have been undertaken specifically because achieving EPHP criteria was challenging, or previous data were difficult to interpret. Nevertheless, our sensitivity analyses did not indicate bias from any single EU, and the included datasets spanned a broad range of prevalence values across the spectrum of trachoma endemicity. As more infection data are collected in future surveys our frameworks can be updated and thresholds reevaluated accordingly if necessary.

Our results demonstrate how by using informative priors, most likely informed by TF_1-9y_ and SCR in this context, a variable infection prevalence threshold may be applied to have the same level of confidence in whether to deliver interventions or not. All metrics, including infection prevalence as measured by PCR, carry specific advantages and limitations both in their implementation and interpretation. For tests requiring laboratory assays there will be a time lag due to sample transport, processing and analyses, as compared to diagnosis of TF which is done in the field, a difference which is of particular relevance when time-critical decisions are necessary. However, change in TF_1-9y_ is generally found to lag behind change in infection after MDA, and a systematic review concluded that TF_1-9y_ may lead to an overestimation of the intensity of transmission in a community after MDA.^38^ SCR in children aged 1–5 years should provide an accurate average of the transmission dynamics over recent years, potentially more representative of average transmission than the single snap-shot that infection prevalence provides, yet may be more insensitive to very recent changes or unusual population dynamics, for example, large scale migration over a short period of time. Defining how multiple population-level metrics can be used concurrently and pragmatically to inform trachoma programme decisions will be a focus for future work.

In the 30 years since the launch of the WHO Alliance for the Global Elimination of Trachoma by 2020 (GET2020) in 1996, the global trachoma programme has made substantial progress towards the ultimate aim of eliminating trachoma as a public health problem. The new target date, 2030, is rapidly approaching. In the current climate of funding challenges, as well as emerging and re-emerging threats to global health driving re-prioritisation of resources, optimising the use of technology to strengthen decision-making is a key priority for programmes. We have defined a framework for delineating the need for population-level interventions based on molecular diagnosis of *Ct*, which could be used in conjuction with current approaches to substantiate intervention and surveillance decisions as we approach trachoma elimination.

Combining data-driven statistical approaches with transmission modelling is broadly generalisable to other disease elimination programmes, particularly where new diagnostics become available, and where pragmatic trade-offs are necessary between cost, timeliness and risk.

## METHODS

### Summary ofdatasets

We identified (see Supplementary Materials for original search criteria) a total of 104 trachoma surveys that had included PCR testing for ocular *Ct* infection. Our inclusion criteria were:

- Commercial PCR *Ct* assay used in analysis
- Availability of either:
  i) Individual-level data,
  ii) Cluster-level numerator/denominator data, or,
  iii) Pool-level results (including individual pool size and pool positive/negative result) for surveys where swabs were pooled.

Of these, five surveys were excluded due to use of a non-commercial PCR assay, and 13 due to insufficient data, leaving 86 surveys for inclusion in our analysis (Supplementary Figure S1). These comprised 68 surveys with individual-level data for children aged 1–5 years, six surveys with cluster-level data for either children aged 1–6 years or 1–5 years, four surveys with cluster-level data for children aged 1–9 years, and eight surveys with pool-level data for children aged 1–9 years.

The final combined dataset used for this analysis represents 60,559 children aged 1–9 years (46,914 from children aged 1–5 years) from 86 surveys carried out between 2007 and 2024 across 13 countries, 12 in Africa: Ethiopia (n=44), Ghana (n=6), Kenya (n=3), Malawi (n=6), Mozambique (n=6), Niger (n=6), Nigeria (n=2), South Sudan (n=1), Sudan (n=1), Uganda (n=5), United Republic of Tanzania (n=4), and Zambia (n=1), plus the Republic of Naoero (n=1) in the South Pacific.

All surveys used quasi-random and/or random sampling and a two-stage cluster design, with a group of households (typically a village) defined as a study cluster. Most surveys (n=83) were representative of a single evaluation unit (EU), with two representing a group of EUs (zonal level survey, Ethiopia-TAITU/Wag Hemra-2018 and Ethiopia WUHA/Wag Hemra-2016), and the Republic of Naoero representing a whole-country survey. An EU is an administrative unit typically representing 100,000–250,000 individuals and is the WHO-recommended population unit in which trachoma activities take place.^25^ Specifics of PCR platforms and assays were described in the original publications (Table 1).

### Expert classification ofprogrammatic categories

Using a secure online survey, we compiled all available summary data comprising EU-level point prevalence estimates of clinical markers of trachoma, specifically TF and trachomatous inflammation—intense (TI) in children aged 1–9 years, anti-Pgp3 seroprevalence and SCR estimates in children aged 1–5 years, and point estimates of *Ct* prevalence (in either children aged 1–5 years, where available, or children aged 1–9 years). In total, 72 individuals identified as having the relevant experience and expertise in trachoma epidemiology and/or diagnostics were invited to complete the survey.

Survey participants were requested to categorise each population as defined below:

**“Action needed”** == “The data suggest that population-level interventions (such as MDA) are needed for this district as it is very likely there is ongoing transmission that could lead to development of disease sequelae and eventually blindness.”

**“No action needed”** == “The data suggest that population-level interventions (such as MDA) are not needed for this district.”

**“I am not sure about this”** == “I don’t know. It is hard to tell where to place this district.”

The survey was completed by 41 individuals (57% response rate). A consensus threshold of 70% agreement (a supermajority) was used to assign categories. EUs without a consensus supermajority were designated “Unclassified”. Using these categorisations and the consensus threshold, our aim was to identify populations in which programmatic decisions would be very clear, allowing for the fact that some populations would fall between the two extremes. We assumed that in these cases, further investigations or context may be necessary to make a decision.

### Prevalence distributions for *Ct* infection by trachoma category

For datasets with individual or cluster-level data, we estimated the EU-level prevalence of *Ct* infection as measured by PCR (*π*), using a Bayesian extension of a standard generalised linear model (GLM) with uninformative priors. Here the number of PCR-positive individuals (*n_pos_c_*) in each cluster (cluster size *N_c_*) was modelled as a binomial response variable,

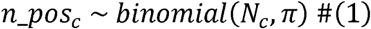

For pool-level datasets, the probability of a positive pool (*pcr_pos_p_*) was modelled as:

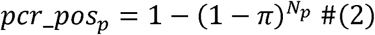

where *N_p_* is pool-size.

Posterior distributions were generated using 8400 draws of a Monte Carlo Markov Chain (MCMC) implemented in Stan (R version 4.5.2).

### Bayesian mixture model and threshold estimation

Posterior prevalence distributions were grouped by expert consensus category (“Action needed” and “No action needed”) to generate category-specific probability density functions. Following Kamau *et al.*,^23^ these pooled probability functions were used to define the likelihoods, *p*(*π*|*θ_k_*); *k∈* {1=“Action needed”, 2=“No action needed”}.

Using Bayes’ rule we then generated a posterior probability for each category *θ_k_* as a function of prevalence *π* (*p*(*θ_k_*|*π*):

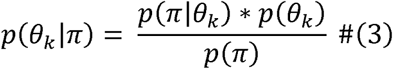

where *p*(*θ_k_*) is the prior probability that a population is in category *θ_k_*. We considered three prior scenarios: *p*(*θ_NO action needed_*) = {0.2,0.5,0.8}; with prior probability of “Action needed” then calculated as 1- *p*(*θ_NO action needed_*) = {0.8,0.5,0.2}. We calculated the marginal likelihood *p*(*π*) by integrating the products of the likelihood *p*(*π*| θ_k_) and the prior.

For all unclassified EUs (n=25) we applied the same Bayesian model to estimate the probability of each programmatic category.

Thresholds for programmatic action were defined using the posterior probability distributions generated from the Bayesian mixture model. We identified prevalence regions that corresponded to ≥90% and ≥95% probability of category *θ_k_*, i.e. either “Action needed” or “No action needed” for each set of priors and interpreted the bounds of these regions as thresholds corresponding to each given level of confidence.

For each unclassified EU (n=25), we compared the median of the prevalence distributions to the 90% uninformative threshold for “No action needed” (0.81%). We estimated the probability that infection prevalence was below this example threshold as the proportion of the posterior distribution that fell below this boundary.

### Impact of timing post-MDA in interpretation of infection prevalence

Observed infection prevalence in a community after a programme of MDA is expected to be relatively sensitive to the time elapsed since last MDA. Given that EU-level decisions ideally need to be made within 12 months after MDA (to allow continuation of annual MDA if indicated), we used a simulation-based modelling approach to explore the dynamics of *Ct* infection after MDA is carried out and the possible implications for interpretation of infection prevalence in the 6–12-month post-MDA TIS window.

### Dynamic transmission model

We used an agent-based model of *Ct* transmission (updated from a previously described trachoma model)^39,40^ implemented in R version 4.5.2, to track prevalence of infection in simulated populations after MDA. The model is detailed in the supplementary materials, and all model code is available on Github.^41^ Briefly, the model is a mechanistic, individual-based stochastic model which explicitly tracks *Ct* infection and TF in individuals. The model incorporates key aspects of ocular *Ct* biology and the clinical course of infection and disease, including TF lagging behind clearance of infection and a decreasing duration of both infection and TF with repeated exposure and increasing age.^20,21^

800 simulations were fitted to each of the pooled posterior empirical distributions of *Ct* infection prevalence for categories “Action needed” and “No action needed” at 12 months post-MDA, generating two model scenarios representing each category with parameter and stochastic uncertainty. Annual population-wide MDA was simulated, with the number of rounds determined by the modelled prevalence of TF_1-9y_ prior to MDA, in accordance with the current guidelines for trachoma programmes.^25^ For pre-MDA TF_1-9y_ of 5–9.9%, one round of MDA was simulated; for TF_1-9y_ of 10–29.9%, three rounds of MDA were simulated; and for TF_1-9y_ ≥30%, five rounds of MDA were simulated. Simulated MDA coverage was between 75 and 90%, and modelled scenarios then tracked transmission with a one-week timestep for up to 13 months after MDA. Model scenarios are illustrated in Supplementary Figure S6.

Prevalence of infection in children aged 1–5 years was extracted from each simulation every week for timesteps between 6 months and 13 months post-MDA (31 timepoints). These were used to generate prevalence probability density distributions at each post-MDA time point for “Action needed” and “No action needed” scenarios, each based on the 800 simulations.

### Estimating thresholds from simulated data

We pooled simulated prevalence estimates for each of the two modelled scenarios “Action needed” or “No action needed” at each post-MDA time point. We then applied the same Bayesian mixture model as for the pooled empirical datasets to the simulated datasets at each post-MDA time point. This generated a time-adjusted posterior probability for each category *θ*_k,t_ as a function of simulated prevalence *π*_s,t_ for each timepoint *t,* post-MDA:

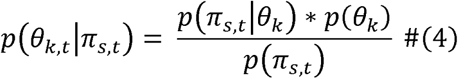

where *t* = {26 *weeks*, 27 *weeks* … 56 *weeks*}. Uninformative priors were used for generating the posterior probabilities. We used the posterior probability distributions generated at each timepoint to estimate time-adjusted prevalence thresholds corresponding to ≥90% and ≥95% probabilities of each category (80% and 85% thresholds also estimated and plotted in Supplementary Materials Figure S9).

### Sensitivity analyses

To assess the potential impact of survey timing relative to MDA, we carried out a series of sensitivity analyses. We subdivided the empirical datasets into those in which the survey was carried out ≥12 months after MDA, and those that were carried out at ≤12, 11, 10 and 9 months after MDA. For each subset of data, we generated the posterior probabilities of the two programmatic categories and estimated the corresponding 90% and 95% probability thresholds.

We also considered the potential for bias using n-1 jack knife resampling sensitivity analyses. Here we excluded each country (n=13) and then each classified EU (n=61) iteratively, estimating the posterior probabilities of the two programmatic categories for each n-1 subset of data, and compared the mean of all leave-one-out iterations to the thresholds calculated for the whole dataset (Supplementary Figures S10 and S11).

There are a number of practical and epidemiological reasons why the focus of our analyses was the infection prevalence in children aged 1–5 years rather than those aged 1–9 years, as has been the case for TF_1-9y_. Briefly, the 1–5 years age group is considered the core group for transmission^18,31^ and it is also considered easier to sample this age group when going house-to-house, as children aged 6–9 years may be in school or (if present) more likely to refuse. Furthermore, focusing on children aged 1–5 years for future surveys will align with trachoma serological survey methodologies.^23^ For surveys where individual-level data were available, we excluded children aged 6–9 years, but this was not possible for those surveys where only cluster- or pool-level data were available. To determine if prevalence estimates were sensitive to inclusion of the 6–9-year-old age group and if prevalence estimates for the 1–5-year-old age group should be age-adjusted, we compared prevalence estimates for children aged 1–9 years and those aged 1–5 years in the individual-level datasets in turn, and compared with age-adjusted prevalence estimates (Supplementary Figures S12 and S13).

## ACKNOWLEDGEMENTS AND FUNDING

This research was supported by The Royal Society (award ref. DHF/R1/221122 to A.B.) and funded in part by the National Institutes of Health (R01AI158884 to B.F.A., R01EY025350 to T.M.L., UG1EY030833 to T.M.L.). M.I.S.D. and S.L.T.C. are staff members of the Pan American Health Organization. A.B.K. and A.W.S. are staff members of the World Health Organization. The authors alone are responsible for the views expressed in this article and they do not necessarily represent the views, decisions or policies of the institutions with which they are affiliated. The Carter Center would like to thank Abbott for its donation of the m2000 RealTime molecular diagnostics system and consumables. The authors would like to thank Diana Martin, Sarah Gwyn and Brook Goodhew for their valued contribution to discussions.

## AUTHOR CONTRIBUTIONS

Conceptualization: A.B., A.W.S., T.M.L., B.F.A., E.K.

Data contribution and/or data curation: R.P-C., K.K.R., F.T.A., P.A., A.A., S.C.A., S.A., Sy.A., Z.A., G.A., M.S.B., R.L.B., S.B., S.Bo., A.Ba., T.B., Y.Y.B., L.B., E.K.C., D.C., B.C., A.C., A.A.D., M.D., D.D., B.E.E., S.E.O., M.F., M.F.S., K.G., T.G., T.A.G., H.E., E.M.H-E., P.J.H., K.A.J., C.Ji., B.K., J.M.K., K.K., H.M.K., M.K., F.K., J.D.K., A.B.K., R.W.K., N.S.K., S.B.L., S.L.B., K.L., B.M., M.P.M., S.G.M., H.M., T.M., C.Mp., K.I.M.M., C.Mw., C.Jo., B.N., S.V.N., J.N., S.M.N., S.O., C.O., N.O., R.O., W.E.O., F.O., M.I.S.D., A.M.S., E.S., F.S., L.G.S., E.A.S., H.M.S., H.S., J.S., Z.T.G., S.L.T.C., E.M.T., T.W-M., T.W.K., S.K.W., D.M.W., D.Y-M., M.Z., T.Z., T.M.L., S.D.N.

Formal analysis: A.B., J.A.C., E.K.

Funding acquisition: A.B., B.F.A.,

Investigation: A.B., J.A.C., E.K.

Methodology: A.B., J.A.C., E.K., B.F.A.

Project administration: R.P-C., K.K.R., A.B., E.K., B.F.A.

Software: E.K., J.A.C., A.B.

Supervision: A.B., A.W.S.

Writing: Original Draft Preparation: A.B., J.A.C., E.K., S.D.N., T.M.L., R.P-C., B.F.A., A.W.S.

Writing: Review and Editing: A.B., J.A.C., E.K., R.P-C., K.K.R., F.T.A., P.A., A.A., S.C.A., S.A., Sy.A., Z.A., G.A., M.S.B., R.L.B., A.Ba., S.B., T.B., Y.Y.B., L.B., S.Bo., E.K.C., D.C., B.C., A.C., A.A.D., M.D., D.D., B.E.E., S.E.O., M.F., M.F.S., K.G., T.G., T.A.G., H.E., E.M.H-E., P.J.H., K.A.J., C.Ji., B.K., J.M.K., K.K., H.M.K., M.K., F.K., J.D.K., A.B.K., R.W.K., N.S.K., S.B.L., S.L.B., K.L., B.M., M.P.M., S.G.M., H.M., T.M., C.Mp., K.I.M.M., C.Mw.,C.Jo., B.N., S.V.N., J.N., S.M.N., S.O., C.O., N.O., R.O., W.E.O., F.O., M.I.S.D., A.M.S., E.S., F.S., L.G.S., E.A.S., H.M.S., H.S., J.S., Z.T.G., S.L.T.C., E.M.T., T.W-M., T.W.K., S.K.W., D.M.W., D.Y-M., M.Z., T.Z., T.M.L., S.D.N., B.F.A., A.W.S.

## ETHICS AND INCLUSION

Data comprised previously collected anonymised data with no individual identifiers that could allow for tracing back to any individual. All studies included followed relevant local research and ethics protocols in accordance with the Declaration of Helsinki, and informed consent was obtained from all participants or their guardian.

The online expert classification survey was completed by invited participants via a secure survey platform, and participant responses were anonymised. The survey format was reviewed and deemed exempt by the Institutional Review Board at the University of California, San Francisco.

Members from each research team that contributed primary data have participated as collaborators, and team members nominated by each study principal investigator included as co-authors. Input from co-authors was incorporated at all stages to represent stakeholder perspectives.

## DATA AVAILABILITY

De-identified data and code sufficient to allow replication of the analyses will be uploaded via Code Ocean and publicly available on publication.

## Competing interests

K.K.R., P.J.H., A.Ba, S.B., and R.P-C. are employees of, and E.M.H-E. receives salary support from, the International Trachoma Initiative, which receives an operating budget and research funds from Pfizer Inc., the manufacturers of Zithromax® (azithromycin).

## Notes

### Author Declarations

The study was reviewed and deemed exempt by the Institutional Review Board at the University of California, San Francisco.

## REFERENCES

1. World Health Organization. Ending the neglect to attain the sustainable development goals - A road map for neglected tropical diseases 2021–2030. https://www.who.int/publications/i/item/9789240010352 (2020).

2. World Health Organization. Validation ofelimination oftrachoma as a public health problem. https://www.who.int/publications/i/item/who-htm-ntd-2016.8 (2016).

3. Lietman, T. M., Oldenburg, C. E. & Keenan, J. D. Trachoma: Time to Talk Eradication. Ophthalmology vol. 127 11–13 (2020).

4. Renneker, K. K., Emerson, P. M., Hooper, P. J. & Ngondi, J. M. Forecasting the elimination of active trachoma: An empirical model. PLoS Negl. Trop. Dis. 16, e0010563 (2022).

5. Srivathsan, A. et al. District-Level Forecast of Achieving Trachoma Elimination as a Public Health Problem By 2030: An Ensemble Modelling Approach. Clin. Infect. Dis. 78, S101–S107 (2024).

6. World Health Organization. WHO validates Timor-Leste’s elimination of trachoma as a public health problem. https://www.who.int/news/item/03-09-2026-who-validates-timor-leste-s-elimination-of-trachoma-as-a-public-health-problem.

7. Solomon, A. W., et al. Trachoma. Nat. Rev. Dis. Prim. 8, 32 (2022).

8. Trachoma Alternative Indicators Study Data review. https://iris.who.int/server/api/core/bitstreams/02275412-3f8d-4ae7-84a9-4d15e41f4d2d/content (2016).

9. Mpyet, C. et al. Use of complementary indicators in trachoma surveys in Nigeria: results of impact surveys in two local government areas. Int. Health (2026) doi: 10.1093/inthealth/ihag071.

10. Mwale, C. et al. Does persistent active trachoma mandate antibiotic mass drug administration? A comparison of prevalence of trachomatous inflammation-follicular with that of conjunctival infection and anti-chlamydial antibodies, Western Province, Zambia. Int. Health 18, 274–281 (2026).

11. Nash, S. D. et al. The Epidemiology of Ocular Chlamydia trachomatis Infection within Districts Persistently Endemic for Trachoma in Amhara, Ethiopia. Am. J. Trop. Med. Hyg. 111, 105–113 (2024).

12. Sanders, A. M. et al. Serological Responses to Trachoma Antigens prior to the Start of Mass Drug Administration: Results from Population-Based Baseline Surveys, North Darfur, Sudan. Am. J. Trop. Med. Hyg. 111, 49–57 (2024).

13. Lynch, K. D. et al. A national survey integrating clinical, laboratory, and WASH data to determine the typology of trachoma in Nauru. PLoS Negl. Trop. Dis. 16, e0010275 (2022).

14. Sun, Y., Dong, S. & Zhou, Q. Laboratory detection methods for Chlamydia trachomatis infection in children: a review. Eur. J. Clin. Microbiol. Infect. Dis. Off. Publ. Eur. Soc. Clin. Microbiol. 45, 41–56 (2026).

15. See, C. W., et al. How reliable are tests for trachoma?--a latent class approach. Invest. Ophthalmol. Vis. Sci. 52, 6133–6137 (2011).

16. Koukounari, A. et al. Using a nonparametric multilevel latent Markov model to evaluate diagnostics for trachoma. Am. J. Epidemiol. 177, 913–922 (2013).

17. Gebresillasie, S. et al. Inter-Rater Agreement between Trachoma Graders: Comparison of Grades Given in Field Conditions versus Grades from Photographic Review. Ophthalmic Epidemiol. 22, 162–169 (2015).

18. Lietman, T. M., et al. Identifying a sufficient core group for trachoma transmission. PLoS Negl. Trop. Dis. 12, e0006478 (2018).

19. House, J. I. et al. Assessment of herd protection against trachoma due to repeated mass antibiotic distributions: a cluster-randomised trial. Lancet (London, England) 373, 1111–1118 (2009).

20. Bailey, R., Duong, T., Carpenter, R., Whittle, H. & Mabey, D. The duration of human ocular Chlamydia trachomatis infection is age dependent. Epidemiol. Infect. 123, 479–486 (1999).

21. Grassly, N. C., Ward, M. E., Ferris, S., Mabey, D. C. & Bailey, R. L. The natural history of trachoma infection and disease in a Gambian cohort with frequent follow-up. PLoS Negl. Trop. Dis. 2, e341 (2008).

22. World Health Organisation Preventative chemotherapy databank. www.who.int/neglected_diseases/preventive_chemotherapy/sch/en.

23. Kamau, E. et al. Characterizing trachoma elimination using serology. Nat. Commun. 16, 5545 (2025).

24. Subtil, F. & Rabilloud, M. Estimating the optimal threshold for a diagnostic biomarker in case of complex biomarker distributions. BMC Med. Inform. Decis. Mak. 14, 53 (2014).

25. Solomon, A., et al. Trachoma Control, A guide for Programme Managers. https://iris.who.int/bitstream/handle/10665/43405/9241546905_eng.pdf?sequence=1 (2006).

26. Blumberg, S. & Lloyd-Smith, J. O. Inference of R(0) and transmission heterogeneity from the size distribution of stuttering chains. PLoS Comput. Biol. 9, e1002993–e1002993 (2013).

27. World Health Organization. Technical consultation on trachoma surveillance. https://apps.who.int/iris/bitstream/handle/10665/174085/WHO_HTM_NTD_2015.02_en.pdf2015?sequence=1 (2014).

28. Solomon, A. W. et al. Quality Assurance and Quality Control in the Global Trachoma Mapping Project. Am. J. Trop. Med. Hyg. 99, 858–863 (2018).

29. Informal consultation on end-game challenges for trachoma elimination. https://apps.who.int/iris/bitstream/handle/10665/363591/9789240048089-eng.pdf (2021).

30. Harding-Esch, E. M. et al. Tropical Data: supporting health ministries worldwide to conduct high-quality trachoma surveys. International health vol. 17 1–3 (2025).

31. Carson, J. et al. Dynamics of trachoma infection in West Africa revealed by a hidden state model. PLoS Comput. Biol. 22, e1014313 (2026).

32. Chidambaram, J. D. et al. Effect of a single mass antibiotic distribution on the prevalence of infectious trachoma. JAMA 295, 1142–1146 (2006).

33. Melese, M. et al. Comparison of annual and biannual mass antibiotic administration for elimination of infectious trachoma. JAMA 299, 778–784 (2008).

34. Gebre, T. et al. Comparison of annual versus twice-yearly mass azithromycin treatment for hyperendemic trachoma in Ethiopia: a cluster-randomised trial. Lancet (London, England) 379, 143–151 (2012).

35. Lakew, T., et al. Reduction and return of infectious trachoma in severely affected communities in Ethiopia. PLoS Negl. Trop. Dis. 3, e376 (2009).

36. Stoller, N. E. et al. Efficacy of latrine promotion on emergence of infection with ocular Chlamydia trachomatis after mass antibiotic treatment: a cluster-randomized trial. Int. Health 3, 75–84 (2011).

37. Lietman, T. M. et al. Frequency of Mass Azithromycin Distribution for Ocular Chlamydia in a Trachoma Endemic Region of Ethiopia: A Cluster Randomized Trial. Am. J. Ophthalmol. 214, 143–150 (2020).

38. Ramadhani, A. M., Derrick, T., Macleod, D., Holland, M. J. & Burton, M. J. The Relationship between Active Trachoma and Ocular Chlamydia trachomatis Infection before and after Mass Antibiotic Treatment. PLoS Negl. Trop. Dis. 10, e0005080 (2016).

39. Borlase, A. et al. Evaluating and mitigating the potential indirect effect of COVID-19 on control programmes for seven neglected tropical diseases: a modelling study. Lancet. Glob. Heal. 10, e1600–e1611 (2022).

40. Borlase, A., et al. Modelling trachoma post 2020: Opportunities for mitigating the impact of COVID-19 and accelerating progress towards elimination. Trans. R. Soc. Trop. Med. Hyg.

41. Borlase, A. Trachoma_Track Github. https://github.com/AnnaMB123/Trachoma_Track (2023).

