## Supplementary for "Defining ocular *Chlamydia trachomatis* infection prevalence thresholds for the trachoma elimination endgame"

### Supplementary Information

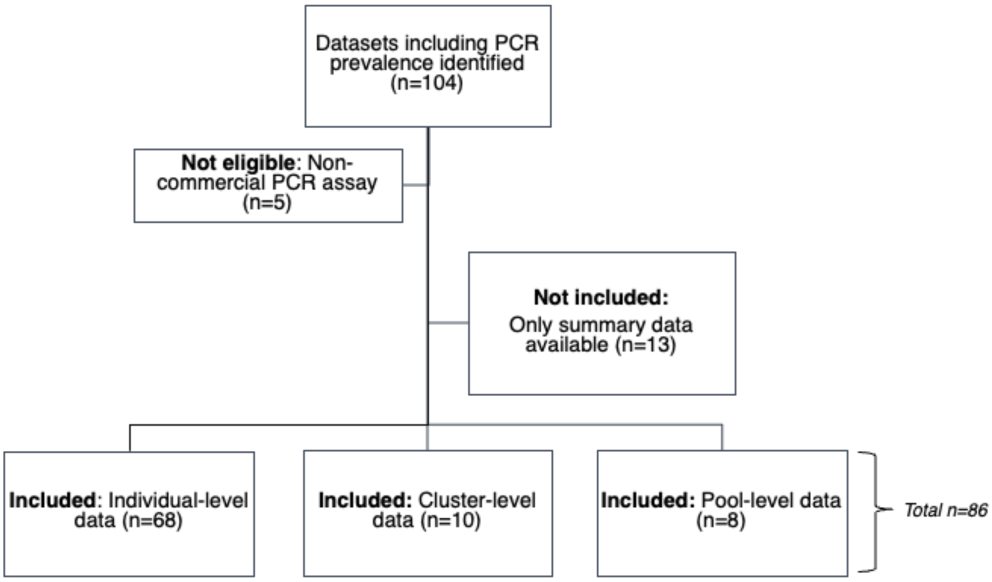


**Supplementary Figure S1. Flow-chart to summarise dataset inclusion criteria.**

Our searches identified 104 population-level trachoma surveys that had included polymerase chain reaction (PCR) testing of conjunctival swabs for *Chlamydia trachomatis* (*Ct*) infection. These searches included a systematic review with protocol registered on PROSPERO (CRD42022356013) and results published previously elsewhere.[1] In addition, through collaboration with the International Trachoma Initiative (ITI), we identified EUs that had included *Ct* infection data as part of their annual reporting to ITI and EUs that had participated in recent multi-country operational research into persistent and recrudescent trachoma, which included testing for *Ct* infection.[2] Finally, through collaboration with The Carter Center, we identified EUs that had included *Ct* infection as part of The Carter Center's surveillance and research activities.

Selection of the final 86 datasets included in our analyses was based on the following criteria:

Use of commercial and externally validated PCR assay and;

Availability of either individual-level, cluster-level (numerator & denominator for each cluster) or pool-level (pool-size and pool-level result) data.

**Supplementary Figure S2. Density distributions of *Ct* infection prevalence (%) for all 86 evaluation units (N=60,559) generated using Bayesian extension of simple generalised linear model (GLM) for binomial data.** Colour represents category “Action needed” (orange; n=30), “No action needed” (blue; n=31) and “Unclassified” (grey; n=25).

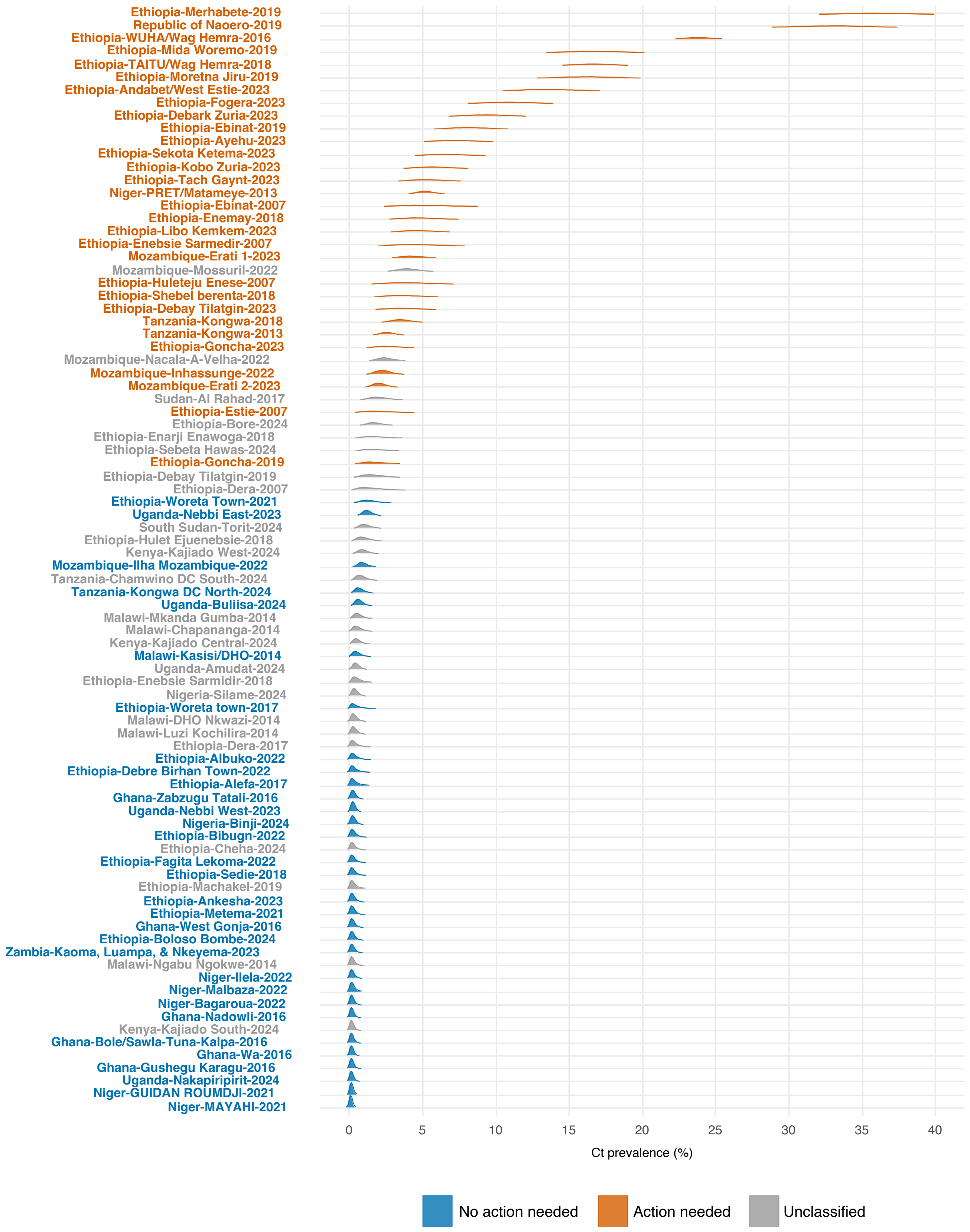
Ethiopia-Merhabete-2019
Republic of Naoero-2019
Ethiopia-WUHA/Wag Hemra-2016
Ethiopia-Mida Woremo-2019
Ethiopia-TAITU/Wag Hemra-2018
Ethiopia-Moretna Jiru-2019
Ethiopia-Andabet/West Estie-2023
Ethiopia-Fogera-2023
Ethiopia-Debark Zuria-2023
Ethiopia-Ebinat-2019
Ethiopia-Ayehu-2023
Ethiopia-Sekota Ketema-2023
Ethiopia-Kobo Zuria-2023
Ethiopia-Tach Gaynt-2023
Niger-PRET/Matameye-2013
Ethiopia-Ebinat-2007
Ethiopia-Enemay-2018
Ethiopia-Libo Kemkem-2023
Ethiopia-Enebsie Sarmedir-2007
Mozambique-Erati 1-2023
Mozambique-Mossuril-2022
Ethiopia-Huleteju Enese-2007
Ethiopia-Shebel berenta-2018
Ethiopia-Debay Tilatgin-2023
Tanzania-Kongwa-2018
Tanzania-Kongwa-2013
Ethiopia-Goncha-2023
Mozambique-Nacala-A-Velha-2022
Mozambique-Inhassunge-2022
Mozambique-Erati 2-2023
Sudan-Al Rahad-2017
Ethiopia-Estie-2007
Ethiopia-Bore-2024
Ethiopia-Enarji Enawoga-2018
Ethiopia-Sebeta Hawas-2024
Ethiopia-Goncha-2019
Ethiopia-Debay Tilatgin-2019
Ethiopia-Dera-2007
Ethiopia-Woreta Town-2021
Uganda-Nebbi East-2023
South Sudan-Torit-2024
Ethiopia-Hulet Ejuenebsie-2018
Kenya-Kajiado West-2024
Mozambique-Ilha Mozambique-2022
Tanzania-Chamwino DC South-2024
Tanzania-Kongwa DC North-2024
Uganda-Buliisa-2024
Malawi-Mkanda Gumba-2014
Malawi-Chapananga-2014
Kenya-Kajiado Central-2024
Malawi-Kasisi/DHO-2014
Uganda-Amudat-2024
Ethiopia-Enebsie Sarmidir-2018
Nigeria-Silame-2024
Ethiopia-Woreta town-2017
Malawi-DHO Nkwazi-2014
Malawi-Luzi Kochilira-2014
Ethiopia-Dera-2017
Ethiopia-Albuko-2022
Ethiopia-Debre Birhan Town-2022
Ethiopia-Alefa-2017
Ghana-Zabzugu Tatali-2016
Uganda-Nebbi West-2023
Nigeria-Binji-2024
Ethiopia-Bibugn-2022
Ethiopia-Cheha-2024
Ethiopia-Fagita Lekoma-2022
Ethiopia-Sedie-2018
Ethiopia-Machakel-2019
Ethiopia-Ankesha-2023
Ethiopia-Metema-2021
Ghana-West Gonja-2016
Ethiopia-Boloso Bombe-2024
Zambia-Kaoma, Luampa, & Nkeyema-2023
Malawi-Ngabu Ngokwe-2014
Niger-Ilela-2022
Niger-Malbaza-2022
Niger-Bagaroua-2022
Ghana-Nadowli-2016
Kenya-Kajiado South-2024
Ghana-Bole/Sawla-Tuna-Kalpa-2016
Ghana-Wa-2016
Ghana-Gushegu Karagu-2016
Uganda-Nakapiripirit-2024
Niger-GUIDAN ROUMDJI-2021
Niger-MAYAHI-2021
0
5
10
15
20
25
30
35
40
Ct prevalence (%)
No action needed
Action needed
Unclassified

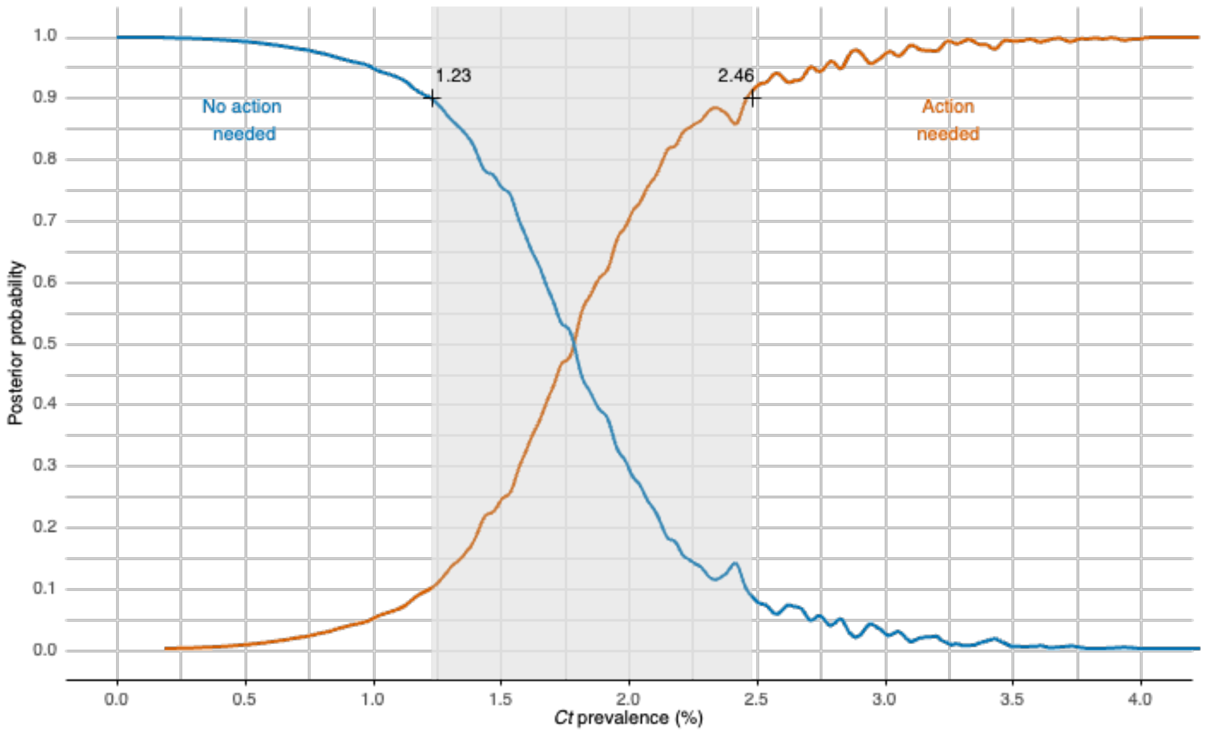


**Supplementary Figure S3. Posterior probabilities of programmatic category ”No action needed” (blue) or "Action needed” (orange) given prevalence of *Ct* infection in children aged 1–5 years. Informative priors have been assumed (80% prior probability for “No action needed”, 20% probability for “Action needed”).**

Prevalence values corresponding to 90% probability thresholds are highlighted with crosses, with shaded area representing the range of prevalence values with less than 90% probability of either category.

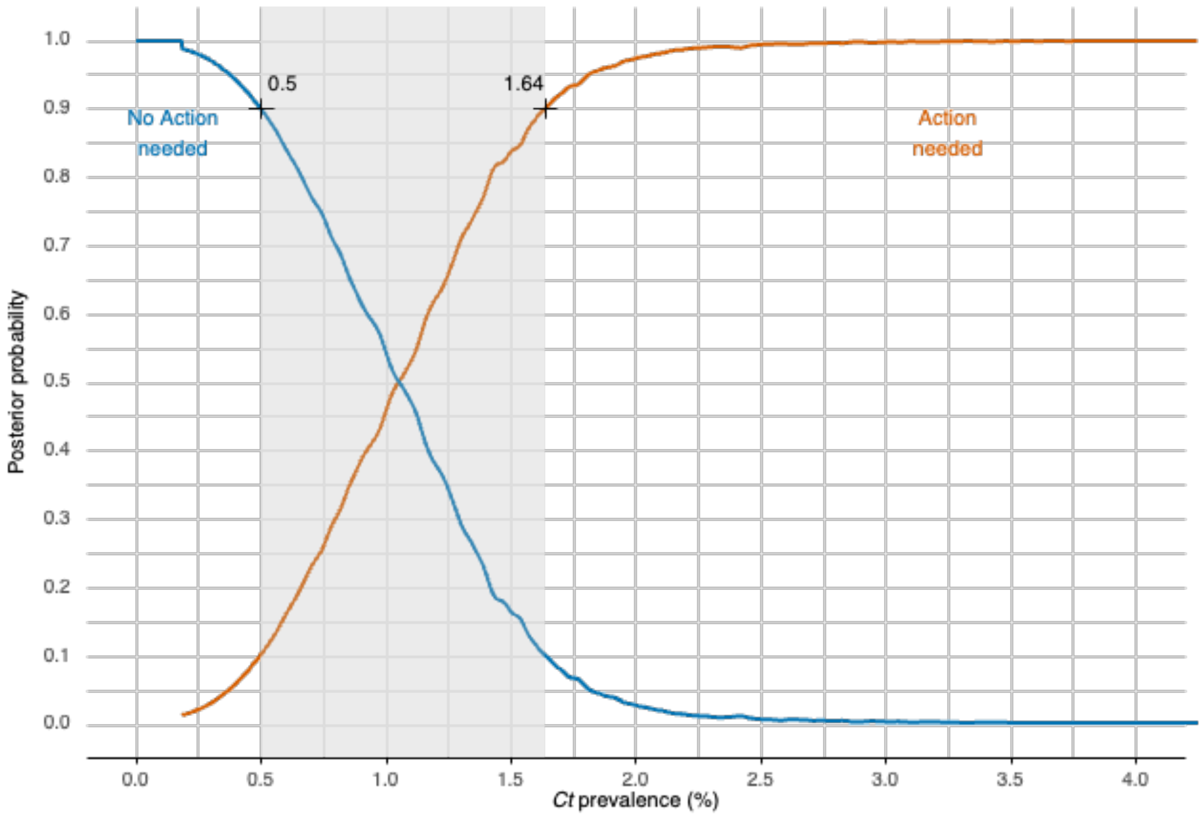


**Supplementary Figure S4. Posterior probabilities of programmatic category "No action needed" (blue) or "Action needed" (orange) given prevalence of *Ct* infection in children aged 1–5 years. Informative priors have been applied (20% prior probability for "No action needed", 80% probability for "Action needed").**

Prevalence values corresponding to 90% probability thresholds are highlighted with crosses, with shaded area representing the range of prevalence values with less than 90% probability of either category.

**Supplementary text: Trachoma transmission model description.**

The dynamic transmission model used is an individual-based (agent-based) stochastic model coded and implemented in R v4.5.2 (https://www.r-project.org) based on previously described models developed under the umbrella of the NTD modelling consortium.[3,4]. All model code is available on GitHub.[5]

Individuals transition through four sequential states illustrated in schematic Supplementary Figure S5: Susceptible (S), infected but not yet diseased (I), infected and diseased (ID) or diseased but no longer infected (D). Here, the diseased state (ID or D periods) represent trachomatous inflammation—follicular (TF) positivity, and infected states (I or ID) are assumed to represent PCR positivity. The model incorporates key aspects of ocular *Ct* infection biology, including acquired immunity leading to decreased duration of infection and disease with repeated infection, and the delayed resolution of TF after infection has been cleared.[6,7] Mass drug administration (MDA) of systemic antibiotic (azithromycin) was simulated with coverage levels between 75 and 90% with probability of clearing infection after treatment at the individual level given as a function of drug efficacy, allowing for simulation of the heterogeneity in infection dynamics after MDA observed in empirical studies.[8,9]

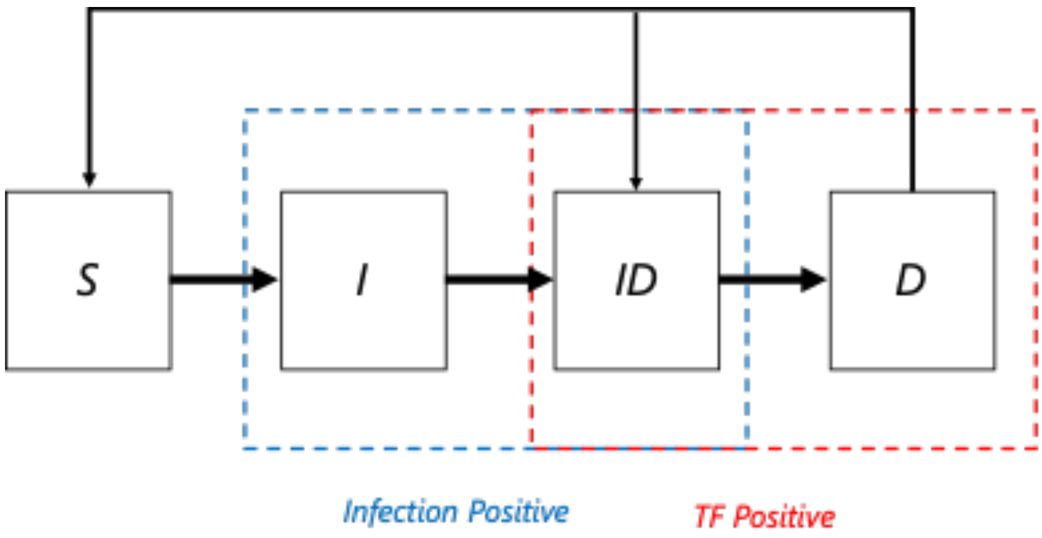


**Supplementary Figure S5. Schematic of agent-based model.**

For each individual *i*, the duration of first ID and D periods ($ID_{i,1}$; $D_{i,1}$) are randomly assigned from Poisson distributions, with distribution means given in Supplementary

Table S3. The duration of these periods for subsequent infections are then assumed to decrease based on both age and number of previous infections with minimum durations and decay rates and given in Table S3. Estimation of these parameters was based on previous analysis of data from the Gambia.[5,6,10] Similarly, it is assumed that an individual's infectivity is proportional to their bacterial load, and that this declines from the first infection following a negative exponential with each subsequent infection (decay parameter given in Table S2). For each individual's ($i$) infection number ($j$), the calculated durations of $ID_{i,j}$ and $D_{i,j}$ are used as fixed transition periods.
Transmission of the ocular strains of $Ct$ is predominantly via close contact and children form the core group for transmission of trachoma.[11,12] Age-disassortative mixing is incorporated into the model, with age of individuals in a simulated population of 2500 people explicitly tracked. We implemented a 200-year burn-in period for all simulations to simulate pre-intervention endemic stability.

Approximate Bayesian Computation was used to fit the model to the empirical data distributions for each programmatic category ("No action needed" or "Action needed"). Number of rounds of MDA to be simulated was determined by TF prevalence in children aged 1–9 years at baseline (post burn-in) for each simulation: For pre-MDA $TF_{1\text{-}9y}$ of 5–9.9%, one round of MDA was simulated; for $TF_{1\text{-}9}$ of 10–29.9%, three rounds of MDA were simulated; and for $TF_{1\text{-}9}$ ≥30%, five rounds of MDA were simulated. Example model simulations are illustrated in Supplementary Figure S6.

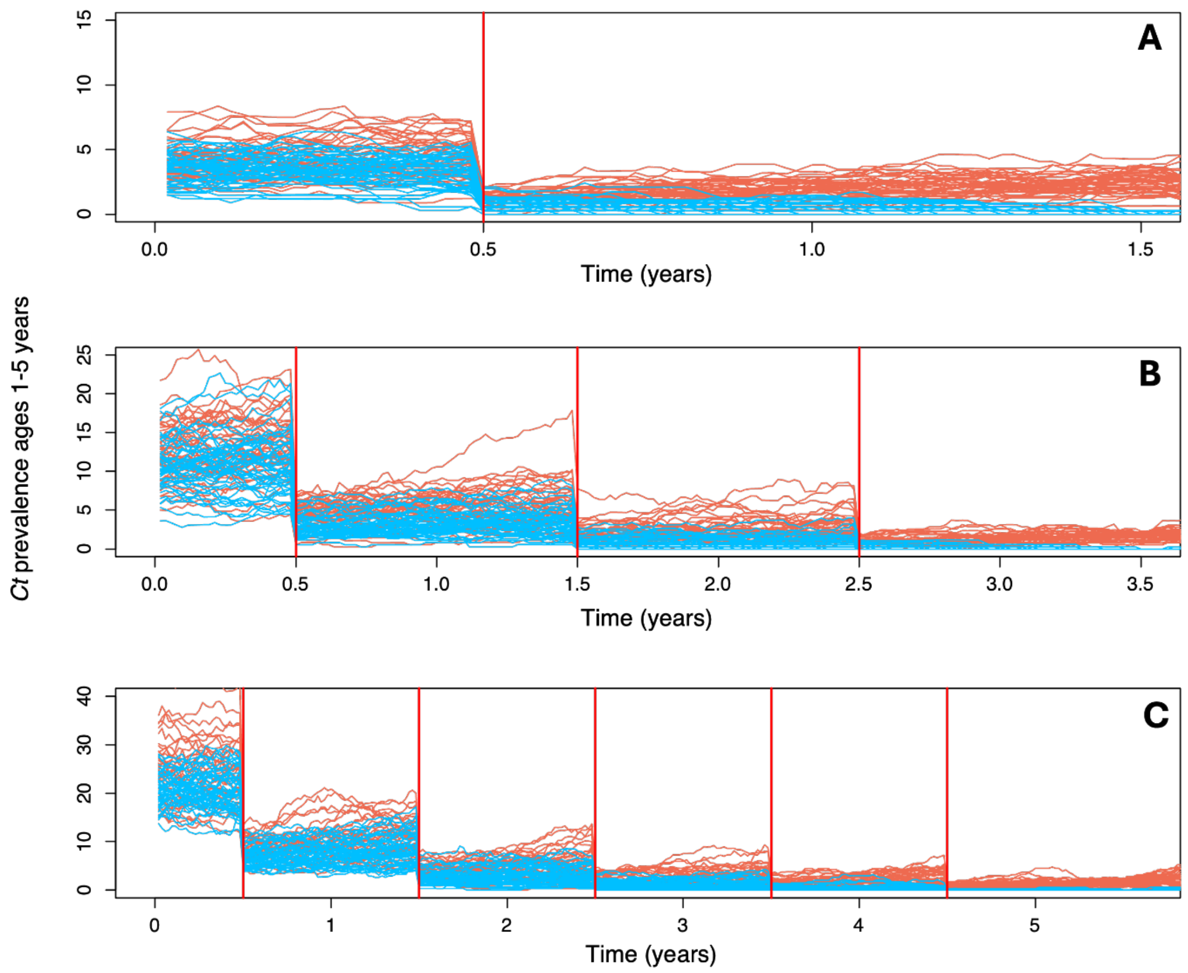


**Supplementary Figure S6. Example of model simulations for A) Hypo-endemic scenario with one round of MDA simulated B) Meso-endemic scenario with 3 rounds of MDA simulated and C) Hyper-endemic scenario with 5 rounds of MDA simulated.** Vertical red lines denote simulated MDA. 800 simulations were fitted to prevalence distributions for "No Action needed" (blue) or "Action needed" (orange) at 12 months post-simulated MDA.

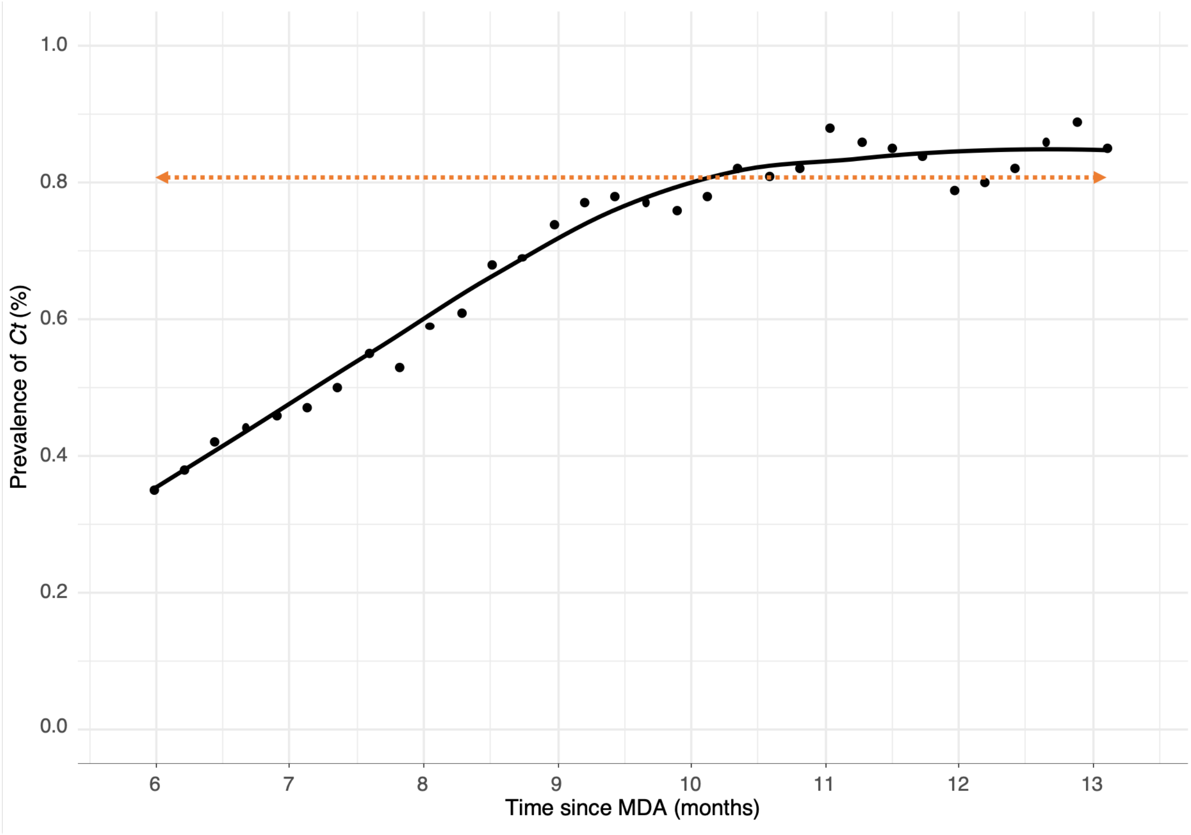


**Supplementary Figure S7. Time adjusted estimates of 90% threshold for "No action needed" based on model-simulated data (black points) and smoothed line generated using Locally Estimated Scatterplot Smoothing (LOESS).** Orange dashed line represents non-time adjusted threshold 0.81% prevalence for more than 90% probability of "No action needed".

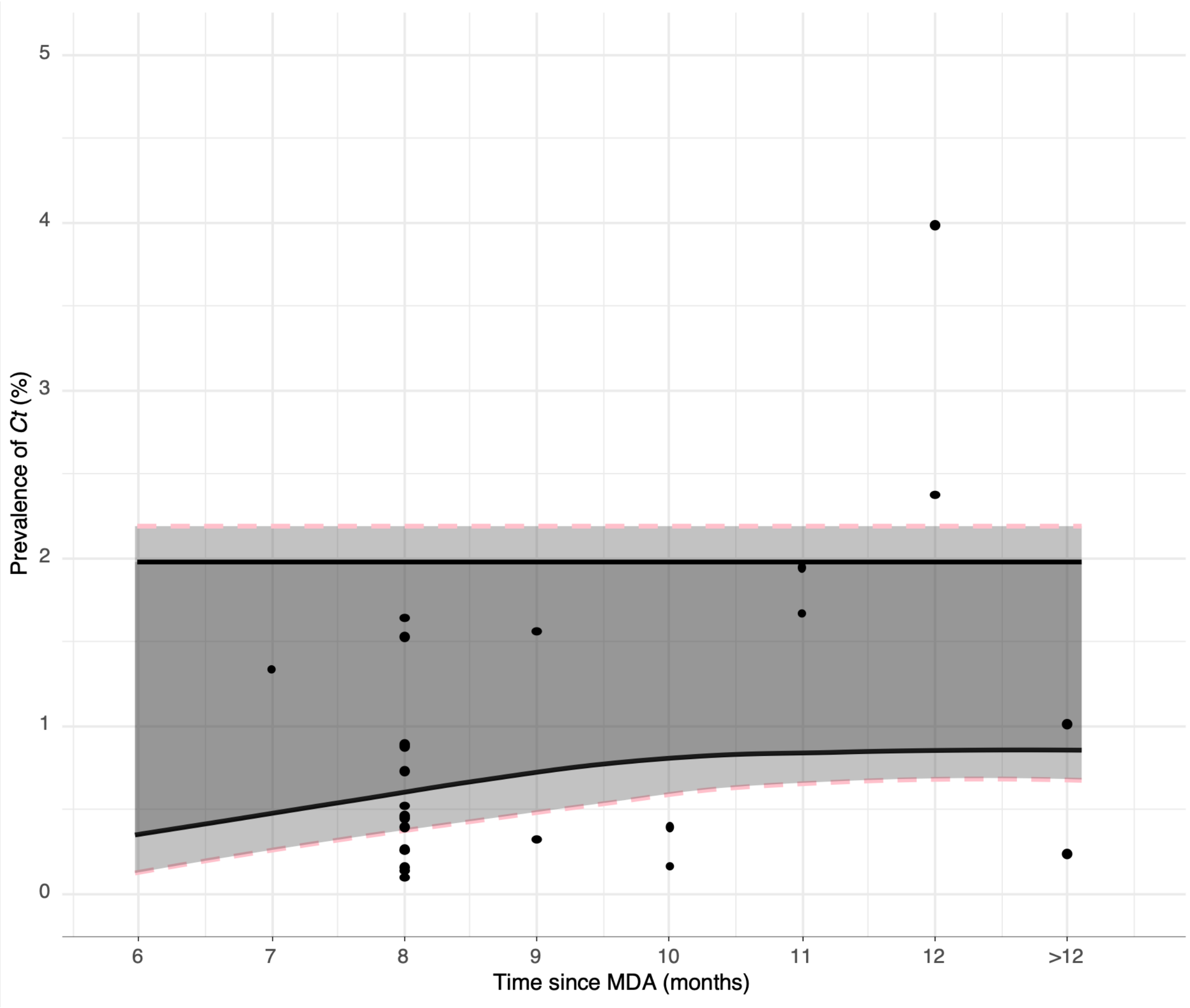


**Supplementary Figure S8. Prevalence point estimates (estimated as median of infection prevalence posterior) for "Unclassified" EUs plotted by time post-MDA compared to proposed time-adjusted 90% programmatic category thresholds (black line) and 95% thresholds (pink dashed line).** The range of prevalence values above the upper pink dashed line is defined as having a 95% or greater probability of programmatic category "Action needed", and the range of prevalence values below the lower pink dashed line corresponds to 95% or greater probability of "No action needed". The light grey regions correspond to a 90–95% probability of "Action needed" (upper light grey region) or "No action needed" (lower light grey region) and the dark grey region corresponds to less than a 90% probability of either category.

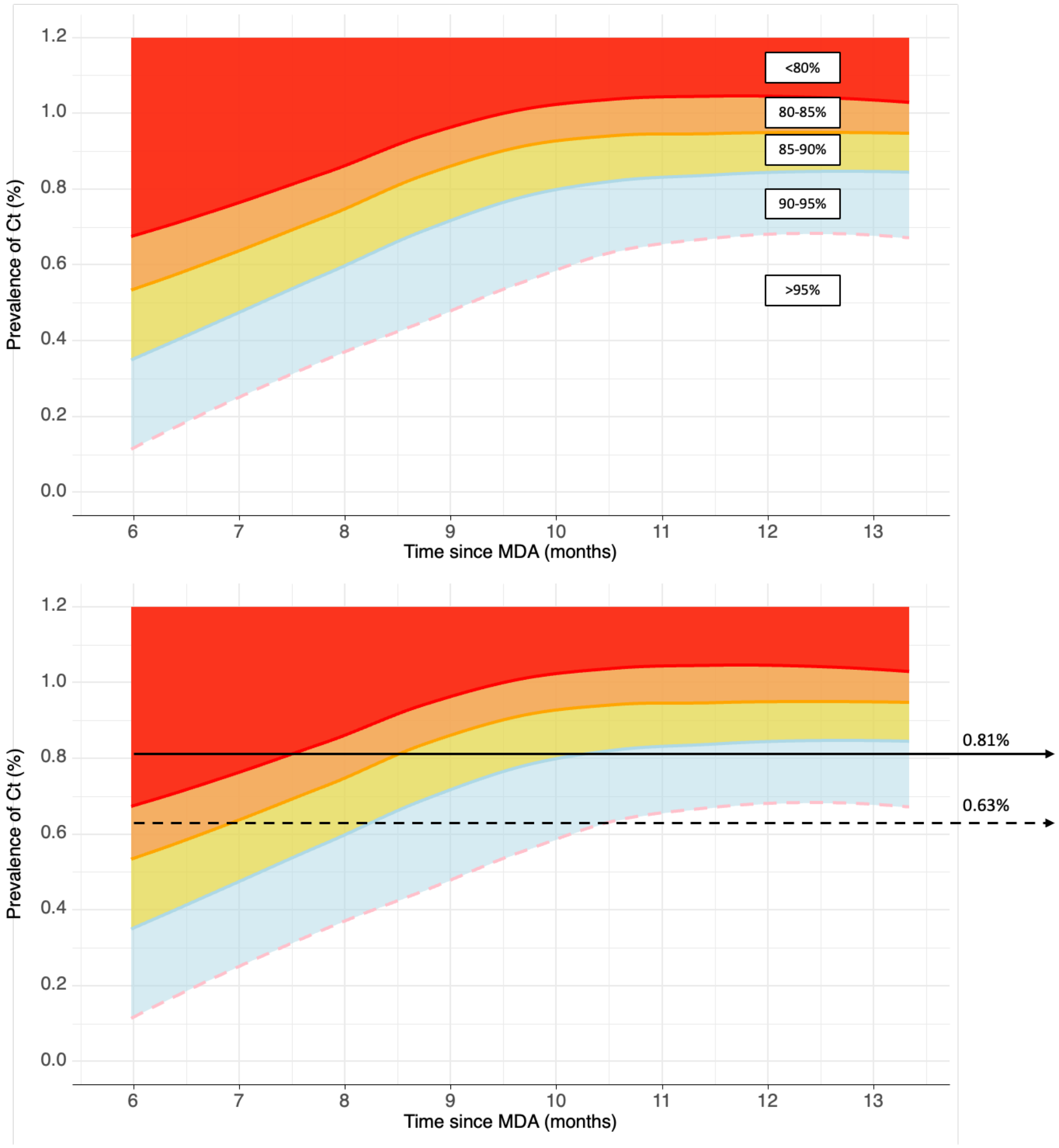


**Supplementary Figure S9. Time-adjusted *Ct* infection prevalence thresholds and ranges for category "No action needed" after a programme of mass drug administration (MDA) generated from model simulations using un-informative priors.** The white area corresponds 95% or greater probability of no action needed, light blue corresponds to 90-95%, yellow 85-90%, orange 80-85%, and red <80%. The solid black line in the lower panel represents the non-adjusted threshold for 90% probability of no action of 0.81%, and the dashed black line represents the non-adjusted 95% threshold of 0.63%. This indicates that by approximately 7 months post-MDA the threshold of 0.63% corresponds to at least an 85% probability of no action needed, and by approximately 8 months this threshold corresponds to at least a 90% probability of category no action needed.

**Supplementary text: Sensitivity analyses.**

The potential for bias from any single country or EU was explored using n-1 "jack-knife" resampling sensitivity analyses, illustrated in Figures S10 and S11. Here we excluded each country (n=13) and then each classified EU (n=61) iteratively, estimating the posterior probabilities of programmatic category for each n-1 subset of data. We assessed the influence of individual countries or populations by comparing the mean of all "leave-one-out" iterations to the thresholds calculated for the whole dataset.

The difference between the thresholds estimated from the full datasets and the mean of the n-1 replicated estimates indicated no bias from any single EU, but that Ethiopia was a highly influential country. The 90% threshold for "No action needed" when Ethiopia is excluded was 1.082% compared to 0.81% for the full dataset. Of the 61 c1assified EUs, 34 were from Ethiopia, and of the 16 highest prevalence datasets (infection prevalence greater than 5%), 14 were from Ethiopia. Ethiopia has the highest burden of trachoma in the world in terms of both number of people in endemic communities and prevalence in those communities. The observed difference in output seen with the inclusion of Ethiopia was therefore considered a representative reflection of the epidemiology and distribution of trachoma.

There are a number of practical and epidemiological reasons why the focus of our analyses was the infection prevalence in children aged 1–5 years rather than those aged 1–9 years, as has been the case for $TF_{1-9}$. Firstly, the 1–5 years age group is considered the core group for transmission and focusing on children aged 1–5 years for future infection prevalence surveys will align with standard serological survey methodologies. An additional factor is that it is generally considered easier to sample children ages 1–5 years in house-to-house surveys, as children aged 6–9 years may be in school or (if present) more likely to refuse. For these reasons prevalence estimates of $TF_{1-9}$ are typically age-adjusted. To determine if prevalence estimates were sensitive to inclusion of the 6–9-year-old age group we compared prevalence estimates for children aged 1–9 years and those aged 1–5 years in the individual-level datasets (Supplementary Figure S12). Within the 1–5 year old age groups we also compared age-adjusted prevalence point estimates to non-adjusted prevalence estimates (Supplementary Figure S13).

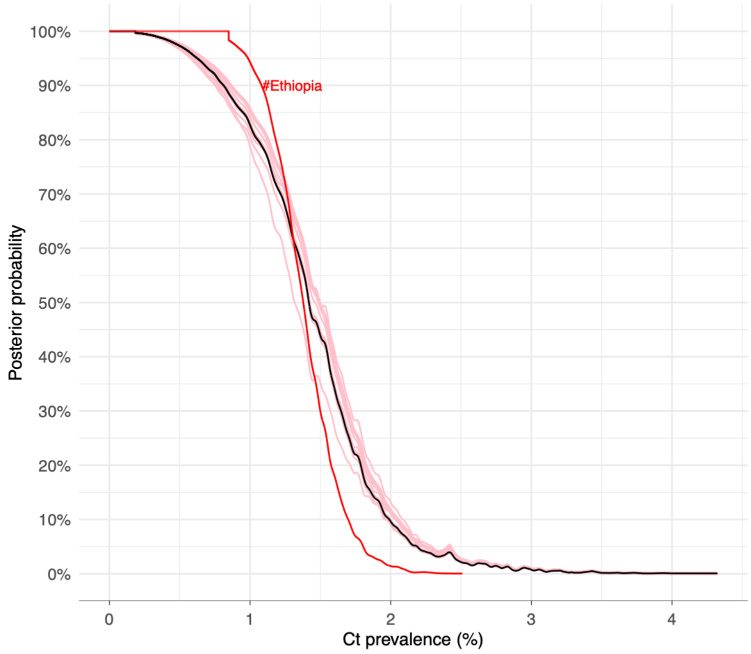


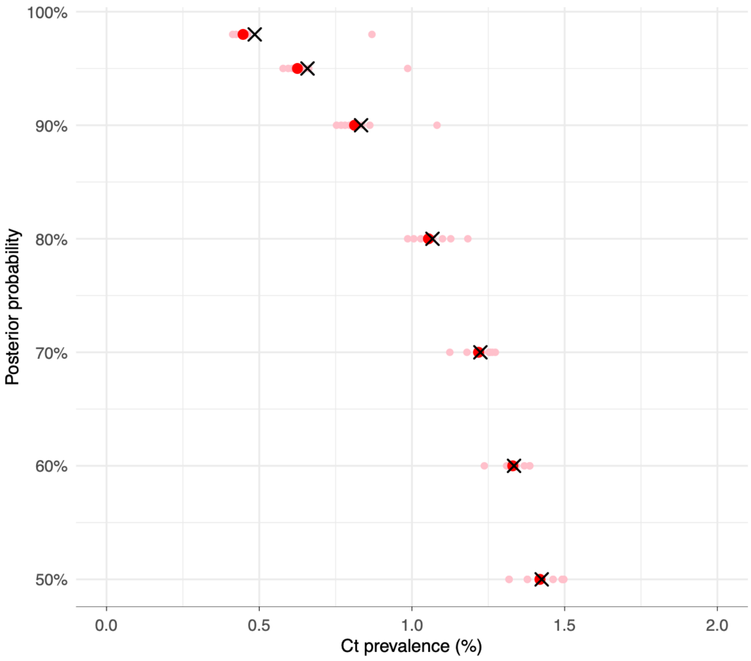


**Supplementary Figure S10. Sensitivity analyses by country.** A resampling approach generating posterior probabilities of programmatic category "No action needed" using n-1 datasets excluding each country in turn. Upper panel: Posterior probability of "No action needed" by *Ct* prevalence (%). Original estimate using datasets from all countries (n=13) shown in black, each n-1 iteration excluding one country in turn in pink. Output for excluding Ethiopia highlighted as potentially highly influential in red. Lower panel: Estimated thresholds for probability of "Action not needed" by *Ct* prevalence (%) for given probabilities (%). Threshold estimates based on all classified datasets (n=61) shown as red circles. Estimates from each n-1 iteration shown as pink circles with mean of these iterations show as a black X.

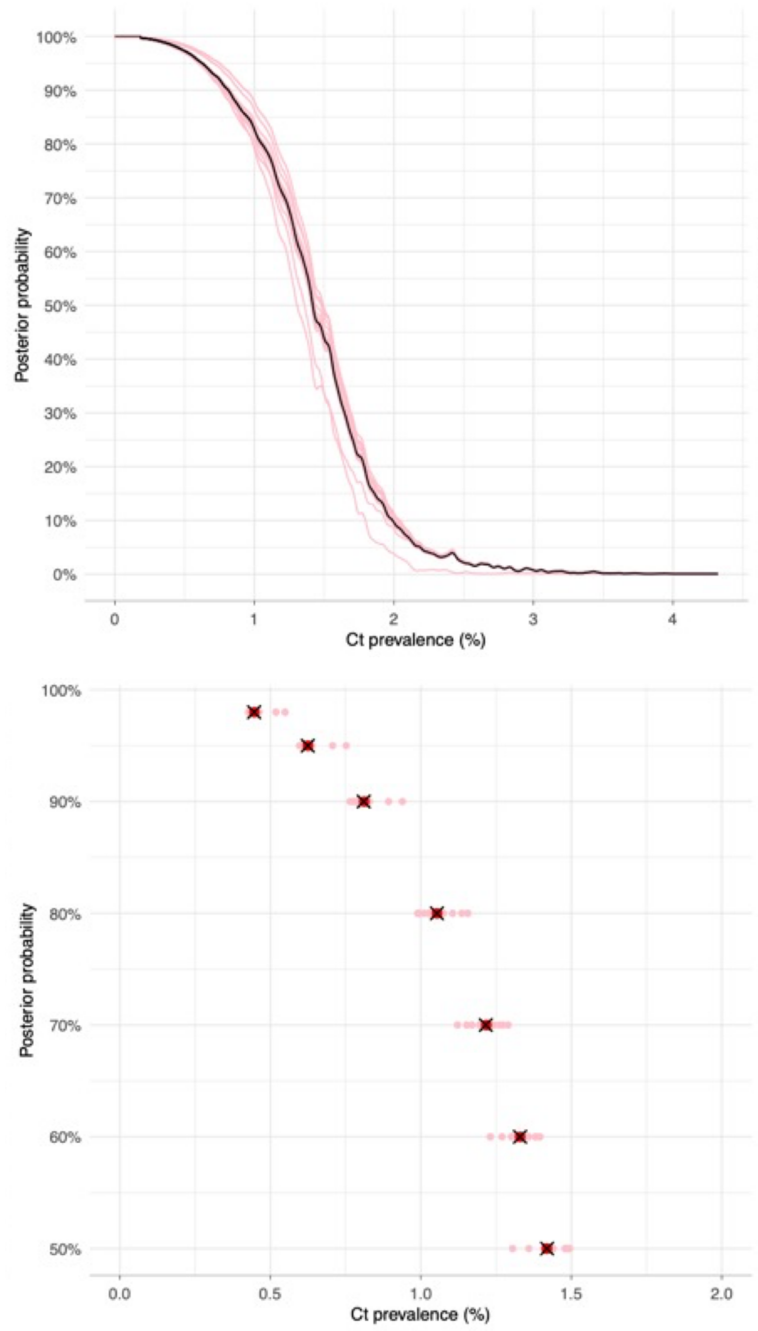


**Supplementary Figure S11. Sensitivity analyses by EU.** A resampling approach generating posterior probabilities of programmatic category "No action needed" using n-1 datasets excluding each EU in turn. Upper panel: Posterior probability of "No action needed" by *Ct* prevalence (%). Original estimate using all classified datasets (n=61) shown in black, each n-1 iteration excluding one EU in turn shown in pink. Lower panel: Estimated thresholds for probability of "No action needed" by *Ct* prevalence (%) for given probabilities (%). Threshold estimates based on all classified datasets (n=61) shown as red circles. Estimates from each n-1 iteration shown as pink circles with mean of these iterations show as a black X.

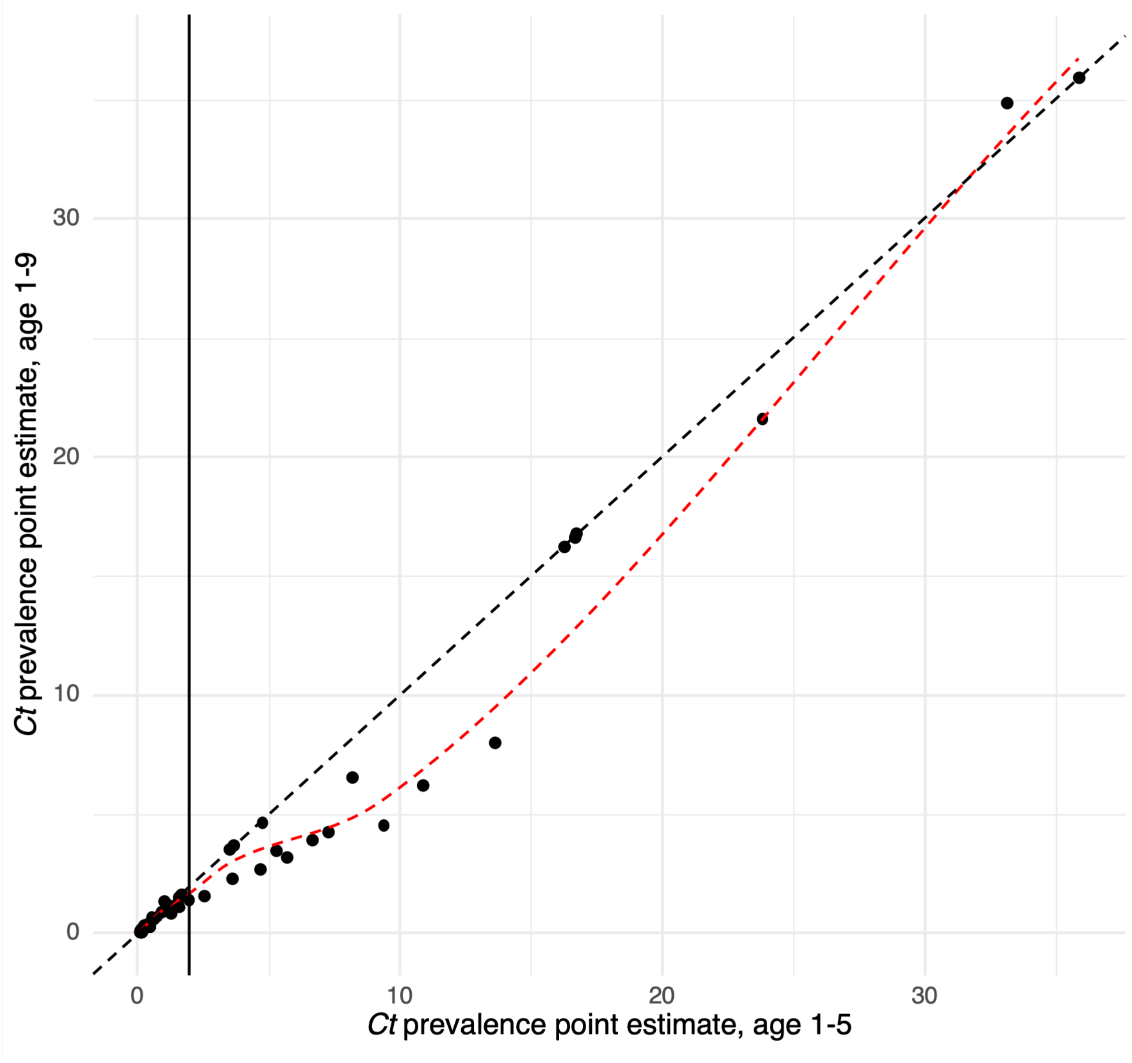


**Supplementary Figure S12. Comparison of *Ct* infection prevalence estimates using different age groups.** For the 68 EUs with individual-level data (n=61,550) a point prevalence estimate was generated as the median of the prevalence posterior for age groups 1–9 years and ages 1–5 years. The 1:1 relationship is plotted as a black dashed line and a locally estimated regression fitted to the prevalence point estimates is shown as a red dashed line. At prevalence levels less that 2.2% (plotted as a black solid line), i.e. within the range of the decision-making thresholds, the prevalence estimated is consistent between the age groups; above this range there is a trend towards lower prevalence when ages 6-9 are included.

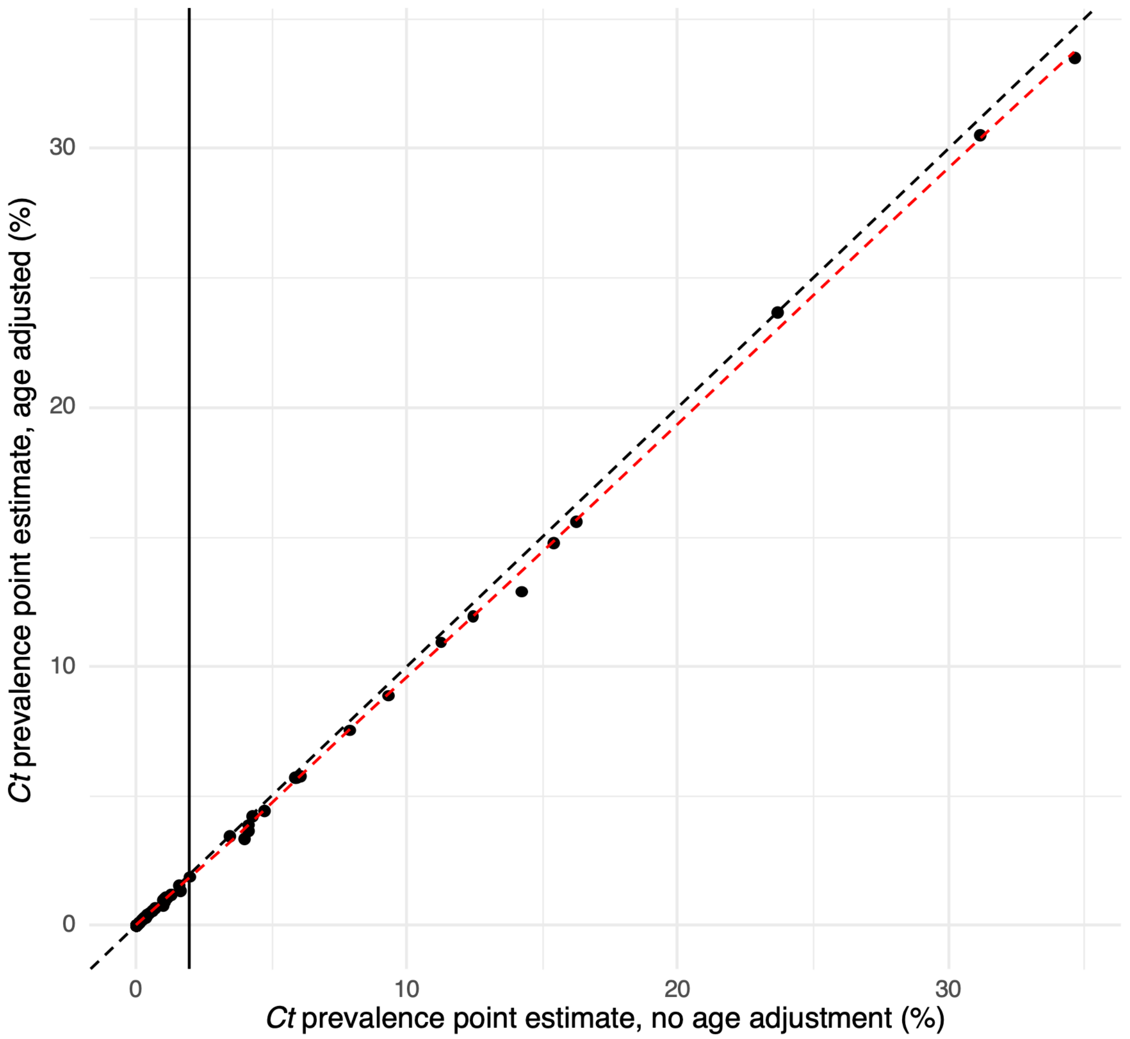


**Supplementary Figure S13. Comparison of age-adjusted and non-age adjusted *Ct* infection prevalence estimates within age group 1–5 years.** For the 68 EUs with individual-level data (n=61,550) a point prevalence estimate was generated as the mean of the cluster-level prevalence, both adjusting for age (assuming uniform distribution of ages between the one and five years old) and not adjusting for age. The 1:1 relationship is plotted as a black dashed line and a locally estimated regression fitted to the prevalence point estimates is shown as a red dashed line. At prevalence levels less that 2.2% (plotted as a black solid line), i.e. within the range of the decision-making thresholds, age-adjusted and non-age adjusted prevalence estimates are consistent.

**Supplementary Text: Estimating time-variable probability of "under-treatment" for proposed thresholds using empirical longitudinal data.**

To assess the potential for time-variable probability of under-treatment (stopping MDA when it is still needed) during the 6–12 months after MDA using our proposed thresholds, we used previously published longitudinal data from the TEF and TANA cluster-randomised trials carried out in Ethiopia and Niger.[8,13–18]

Summary data from these trials included point estimates of *Ct* infection prevalence in children aged 1–9 years old, and we identified communities that had received MDA at baseline and were then followed for at least a year (up to 30 months) without additional rounds of MDA. For 55 communities, infection was observed to be above 1.99% (our proposed threshold for programmatic action being needed with 90% probability) at the end of the observation period. We used the proportion of these communities where the estimated prevalence was below the proposed thresholds for no programmatic action of 0.63% and 0.81% as an approximation for the probability of under-treatment at different time points. For 33 of the 55 communities repeat swabs were taken at 6 months post-MDA and infection prevalence was reported to be below both the 0.63% and 0.81% threshold in 5 of these communities, giving an estimated probability of under-treatment of 15%. At 12 months post-MDA all 55 communities had repeat swabs and reported prevalence was less than the proposed thresholds in 2 communities, giving an estimated probability of under-treatment of 3.6%. By the 24 and 30 month follow-up surveys all communities had reported prevalence greater than the proposed thresholds.

**Supplementary Text: Comparing methods for point prevalence estimation**

When infection surveys for *Chlamydia trachomatis* are carried out there are a number of different methods for calculating a point prevalence estimate. Our proposed thresholds were generated applying a Bayesian extension of Generalised linear model (GLM) for binomial data:

$$n\_pos_c \sim binomial(N_c,\ \pi) \qquad \text{(S1)}$$

Here the number of PCR-positive individuals ($n\_pos_c$) in each cluster (cluster size $N_c$) was modelled as a binomial response variable, with EU-level prevalence then given as $\pi$. When estimating the probability of over-and under-treatment we used the median of the prevalence posteriors for each EU as the point prevalence to be compared to our proposed thresholds.

In order to compare how alternative methods for generating prevalence point estimates may affect EU categorisation, we estimated *Ct* prevalence using two alternative methods for two-stage cluster sampling prevalence surveys:

1) GLM for binomial response using maximum likelihood estimation (MLE) (equivalent to equation S1 above but using MLE).

2) Mean of cluster-level proportion of positives, given in equation S2 (equivalent to method currently used by Tropical Data to estimate TF prevalence in children aged 1–9 years old):

$$\pi = \frac{1}{n}\sum_{i=1}^{n}\frac{n\_pos_{c,i}}{N_{c,i}}$$

Where $n$ is total number of clusters.

For the 55 datasets that were assigned categories based on the expert survey and where either individual-level or cluster-level data were available, we estimated *Ct* prevalence using these two alternative methods. We then compared these to the median of the Bayesian prevalence posteriors and to the categories assigned using expert consensus.

Using all three prevalence estimation methods, no EUs categorised as “Action needed” would be assigned “No Action needed” and vice versa based on our proposed 90% probability thresholds (Supplementary Figures S14 and S15). For all three prevalence estimation methods the same four EUs fall within the intermediate range corresponding to less than 90% probability for either category. The regression fit indicates a slight trend towards the Bayesian posterior median overestimating prevalence as compared to the Tropical Data method (Figure S14), but this does not lead to differences in categorisation. The regression fit indicates almost perfect agreement between the Bayesian posterior median and the MLE estimation of prevalence (Figure S15).

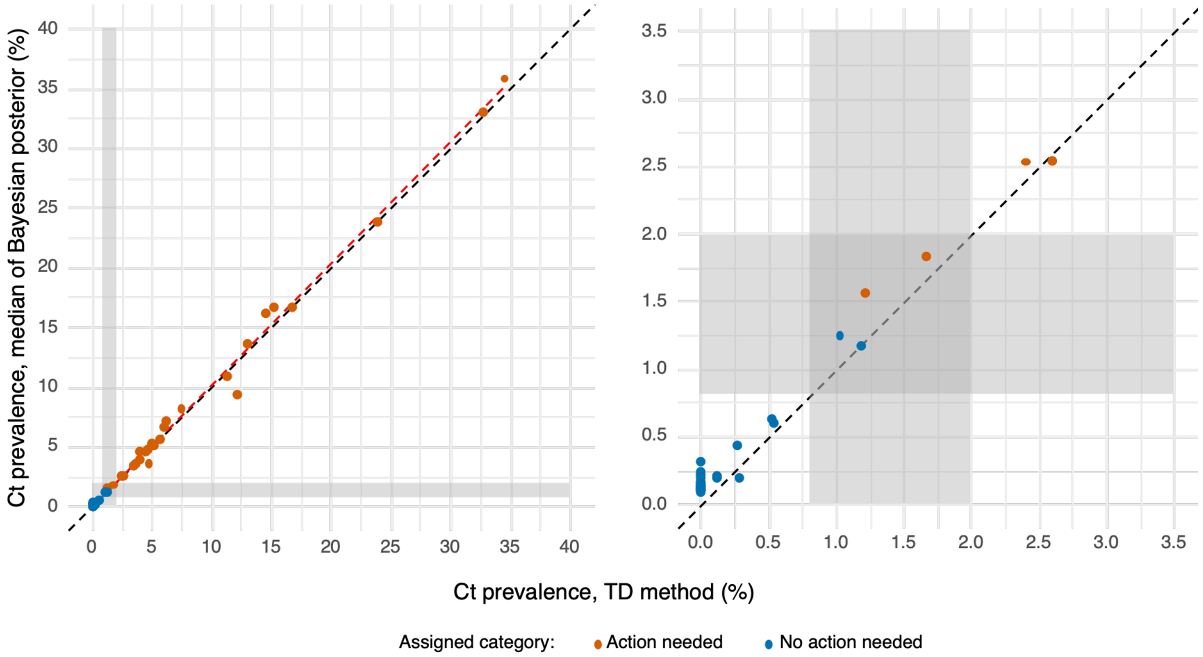


**Supplementary figure S14. Prevalence of *Ct* estimated using median of Bayesian posterior compared to prevalence estimated as mean of cluster proportions (Tropical Data method).**

Left panel: For the 55 EUs (n=36,828) with individual-level or cluster-level data and a consensus categorisation by expert survey, a point prevalence estimate was generated as the mean of the cluster-level proportion of positives (x-axis, "TD method)" and as the median of the prevalence posterior generated using Bayesian extension of GLM (y-axis). The 1:1 relationship is plotted as a black dashed line and a locally estimated regression fitted to the prevalence point estimates is shown as a red dashed line. The grey boxes represent the bounds of the proposed 90% probability thresholds for "No action needed" (0.81% prevalence) and "Action needed" (1.99% prevalence). Colour indicates categorisation by expert survey. Right panel represents the same plot focusing on the lower prevalence range close to the proposed threshold boundaries.

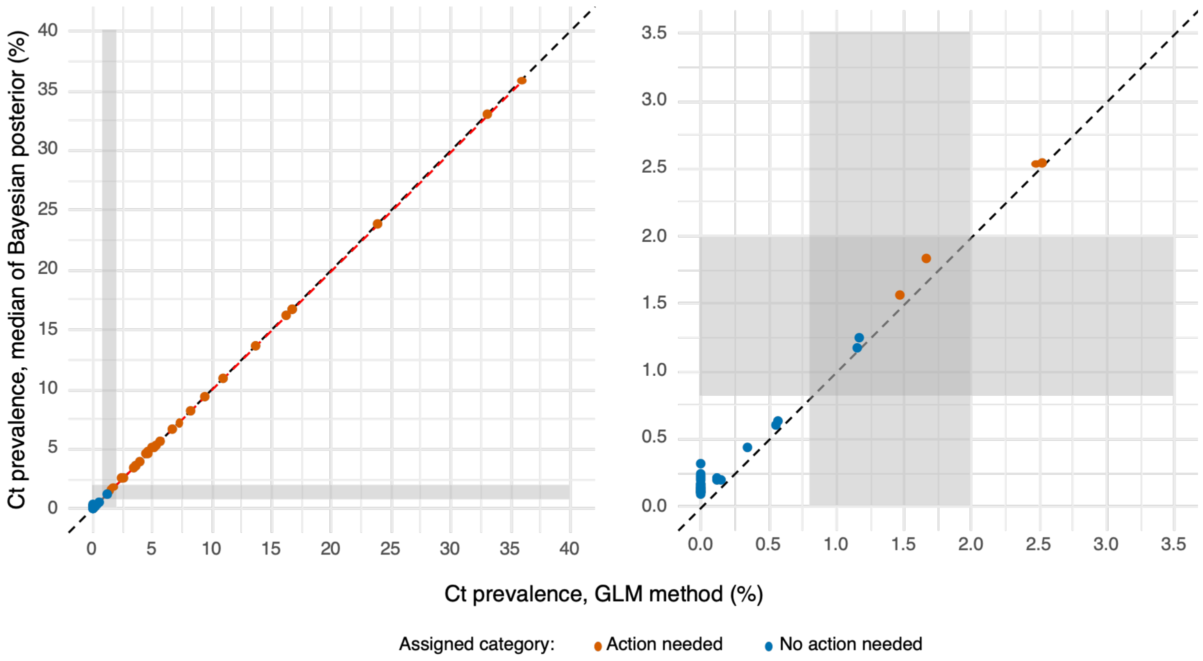


**Supplementary Figure S15. Prevalence of *Ct* estimated using median of Bayesian posterior compared to prevalence estimated using GLM with MLE.**

Left panel: For the 55 EUs (n=36,828) with individual-level or cluster-level data and a consensus categorisation by expert survey a point prevalence estimate was generated using GLM with MLE (x-axis, "GLM method") and as the median of the prevalence posterior generated using Bayesian extension of GLM (y-axis). The 1:1 relationship is plotted as a black dashed line and a locally estimated regression fitted to the prevalence point estimates is shown as a red dashed line. The grey boxes represent the bounds of the proposed 90% probability thresholds for "No action needed" (0.81% prevalence) and "Action needed" (1.99% prevalence). Colour indicates categorisation by expert survey. Right panel represents the same plot focusing on the lower prevalence range close to the proposed threshold boundaries.

#### Supplementary Information Tables

**Supplementary Table S1. Sensitivity analysis of *Ct* prevalence thresholds that correspond to 90% or 95% probability of category "Action needed" or "No action needed" according to timing of survey post-MDA.** An uninformative prior probability for each category was applied. Total number of surveys for each time grouping and number in each category used for calculating given in footnote.

| | **90%** | | **95%** | |
|---|---|---|---|---|
| **≥12 months[1]** | No action needed threshold (%) | Action needed threshold (%) | No action needed threshold (%) | Action needed threshold (%) |
| | 1.078 | 1.996 | 0.944 | 2.431 |
| | **90%** | | **95%** | |
| **≤12 months[2]** | No action needed threshold (%) | Action needed threshold (%) | No action needed threshold (%) | Action needed threshold (%) |
| | 0.883 | 1.724 | 0.706 | 1.823 |
| | **90%** | | **95%** | |
| **≤11 months[3]** | No action needed threshold (%) | Action needed threshold (%) | No action needed threshold (%) | Action needed threshold (%) |
| | 0.802 | 1.649 | 0.623 | 1.797 |
| | **90%** | | **95%** | |
| **≤10 months[4]** | Action not needed threshold (%) | Action needed threshold (%) | Action not needed threshold (%) | Action needed threshold (%) |
| | 0.809 | 1.63 | 0.629 | 1.776 |
| | **90%** | | **95%** | |
| **≤9 months[5]** | No action needed threshold (%) | Action needed threshold (%) | No action needed threshold (%) | Action needed threshold (%) |
| | 0.792 | 1.649 | 0.608 | 1.823 |

**[1]N=36, 10 Action needed, 22 No action needed, 4 Unclassified**
**[2]N=52, 19 Action needed, 10 No action needed, 23 Unclassified**
**[3]N=46, 16 Action needed, 9 No action needed, 21 Unclassified**
**[4]N=43, 16 Action need, 8 No action needed, 19 Unclassified**
**[5]N=37, 15 Action needed, 5 No action needed, 17 Unclassified**

**Supplementary Table S2.**
Summary of Unclassified surveys. Median of *Ct* infection prevalence posterior (children aged 1–5) given with summary expert survey responses for each category as a percentage of all respondents who selected each category "Action needed", "No action needed" or "Unsure". Posterior probabilities of each category generated by applying Bayesian mixture method to prevalence posterior for each EU.

| | | Expert survey responses (%) | | | Posterior probabilities (%) | |
|---|---|---|---|---|---|---|
| **EU** | **Median Ct prevalence (%)** | **Action needed** | **No action needed** | **Unsure** | **Action needed** | **No action needed** |
| Mozambique-Mossuril-2022 | 3.99 | 66.67 | 9.52 | 23.81 | 99.85 | 0.15 |
| Mozambique-Nacala-A-Velha-2022 | 2.38 | 61.90 | 14.29 | 23.81 | 92.80 | 7.20 |
| Sudan-Al Rahad-2017 | 1.95 | 45.24 | 19.05 | 35.71 | 76.36 | 23.64 |
| Ethiopia-Bore-2024 | 1.67 | 45.24 | 28.57 | 26.19 | 65.71 | 34.29 |
| Ethiopia-Enarji Enawoga-2018 | 1.64 | 64.29 | 9.52 | 26.19 | 60.90 | 39.10 |
| Ethiopia-Sebeta Hawas-2024 | 1.57 | 54.76 | 19.05 | 26.19 | 58.26 | 41.74 |
| Ethiopia-Debay Tilatgin-2019 | 1.53 | 50.00 | 21.43 | 28.57 | 56.23 | 43.77 |
| Ethiopia-Dera-2007 | 1.34 | 59.52 | 11.90 | 28.57 | 47.92 | 52.08 |
| South Sudan-Torit-2024 | 1.01 | 55.56 | 11.11 | 33.33 | 24.73 | 75.27 |
| Ethiopia-Hulet Ejuenebsie-2018 | 0.89 | 66.67 | 7.14 | 26.19 | 21.32 | 78.68 |
| Kenya-Kajiado West-2024 | 0.88 | 57.14 | 21.43 | 21.43 | 17.77 | 82.23 |
| Tanzania-Chamwino DC South-2024 | 0.73 | 16.67 | 54.76 | 28.57 | 12.91 | 87.09 |
| Malawi-Mkanda Gumba-2014 | 0.53 | 30.95 | 45.24 | 23.81 | 6.29 | 93.71 |
| Malawi-Chapananga-2014 | 0.47 | 9.52 | 66.67 | 23.81 | 5.10 | 94.90 |
| Kenya-Kajiado Central-2024 | 0.45 | 57.14 | 19.05 | 23.81 | 4.11 | 95.89 |
| Ethiopia-Enebsie Sarmidir-2018 | 0.4 | 38.10 | 26.19 | 35.71 | 5.08 | 94.92 |
| Uganda-Amudat-2024 | 0.4 | 11.90 | 69.05 | 19.05 | 3.02 | 96.98 |
| Nigeria-Silame-2024 | 0.32 | 19.05 | 64.29 | 16.67 | 2.06 | 97.94 |
| Malawi-DHO Nkwazi-2014 | 0.27 | 19.05 | 66.67 | 14.29 | 1.73 | 98.27 |
| Malawi-Luzi Kochilira-2014 | 0.26 | 21.43 | 61.90 | 16.67 | 1.65 | 98.35 |
| Ethiopia-Dera-2017 | 0.24 | 38.10 | 30.95 | 30.95 | 3.08 | 96.92 |
| Ethiopia-Cheha-2024 | 0.17 | 16.67 | 61.90 | 21.43 | 1.26 | 98.74 |
| Ethiopia-Machakel-2019 | 0.16 | 16.67 | 57.14 | 26.19 | 1.26 | 98.74 |
| Malawi-Ngabu Ngokwe-2014 | 0.14 | 19.05 | 52.38 | 28.57 | 0.76 | 99.24 |
| Kenya-Kajiado South-2024 | 0.1 | 23.81 | 45.24 | 30.95 | 0.37 | 99.63 |

#### Supplementary Table S3. Agent-based dynamic transmission model variables, parameters, and sources.

| Notation | Description | Value/distribution | Units | Source |
|---|---|---|---|---|
| $S_i$ | Susceptible individuals | | | |
| $I_{i,j}$ | Infected and not yet diseased (individual *i* infection *j*) | 2 | Weeks | [10,19] |
| $ID_{i,1}$ | Infected and diseased period (individual *i*, first infection) | $\sim Poisson(\omega_{max})$ | Weeks | |
| $ID_{i,j}$ | Infected and diseased period (individual *i*, infection *j*, age=$A_i$) | $(ID_{i,1} - \omega_{min})e^{\phi(j-1)+\gamma A_i} + \omega_{min}$ | Weeks | |
| $D_{i,1}$ | Diseased only period (individual *i*, infection *j*) | $\sim Poisson(\tau_{max})$ | Weeks | |
| $D_{i,j}$ | Diseased only period (individual *i*, infection *j*) | $(D_{i,1} - \tau_{min})e^{\theta(j-1)+\delta A_i} + \tau_{min}$ | Weeks | [6,10] |
| $\omega$ | Duration of ID period | $\omega_{max} = 28, \omega_{min}$=2 | Weeks | [6,10] |
| $\tau$ | Duration of diseased only period | $\tau_{max} = 43, \tau_{min}$=1 | Weeks | [5–7,10] |
| $\phi$ | Decrease in ID period accounted for by previous infections | 0.039 | | [5–7,10] |
| $\gamma$ | Decrease in ID period accounted for by age of individual | 0.00012 | | [5–7,10] |
| $\theta$ | Decrease in D period accounted for by previous infections | 0.078 | | [5–7,10] |
| $\delta$ | Decrease in D period accounted for by age of individual | 0.00098 | | [5–7,10] |
| $b$ | Infectivity of an individual proportional to their bacterial load | 0.114 | Proportion | [10,19] |
| $\beta$ | Transmission parameter | Varied to fit infection prevalence data | | |
| $\epsilon_{MDA}$ | Azithromycin efficacy (probability of infection clearance given that treatment is received) | 0.85 | Proportion | [20] |

#### Supplementary references


1. Renneker KK, Lin C-C, Hsieh JL, et al. Comparison of Methods for Assessing Chlamydia trachomatis Transmission Intensity: A Systematic Review. *Am J Trop Med Hyg*. Published online September 2026. doi:10.4269/ajtmh.25-0541
2. *Persistent and Recrudescent Trachoma Meeting*; 2025. https://www.cor-ntd.org/resources/persistent-and-recrudescent-trachoma-technical-meeting-report
3. Borlase A, Blumberg S, Callahan EK, et al. Modelling trachoma post 2020: Opportunities for mitigating the impact of COVID-19 and accelerating progress towards elimination. *Trans R Soc Trop Med Hyg*.
4. Pinsent A, Hollingsworth TD. Optimising sampling regimes and data collection to inform surveillance for trachoma control. *PLoS Negl Trop Dis*. 2018;12(10):e0006531. doi:10.1371/journal.pntd.0006531
5. Borlase A. Trachoma_Track Github. Published 2023. https://github.com/AnnaMB123/Trachoma_Track
6. Bailey R, Duong T, Carpenter R, Whittle H, Mabey D. The duration of human ocular Chlamydia trachomatis infection is age dependent. *Epidemiol Infect*. 1999;123(3):479-486. doi:10.1017/s0950268899003076
7. Bailey RL, Arullendran P, Whittle HC, Mabey DC. Randomised controlled trial of single-dose azithromycin in treatment of trachoma. *Lancet (London, England)*. 1993;342(8869):453-456. doi:10.1016/0140-6736(93)91591-9
8. Lakew T, House J, Hong KC, et al. Reduction and return of infectious trachoma in severely affected communities in Ethiopia. *PLoS Negl Trop Dis*. 2009;3(2):e376. doi:10.1371/journal.pntd.0000376
9. Oldenburg CE, Amza A, Kadri B, et al. Comparison of Mass Azithromycin Coverage Targets of Children in Niger: A Cluster-Randomized Trachoma Trial. *Am J Trop Med Hyg*. 2018;98(2):389-395. doi:10.4269/ajtmh.17-0501
10. Grassly NC, Ward ME, Ferris S, Mabey DC, Bailey RL. The natural history of trachoma infection and disease in a Gambian cohort with frequent follow-up. *PLoS Negl Trop Dis*. 2008;2(12):e341. doi:10.1371/journal.pntd.0000341
11. Solomon AW, Holland MJ, Burton MJ, et al. Strategies for control of trachoma: observational study with quantitative PCR. *Lancet (London, England)*. 2003;362(9379):198-204. doi:10.1016/S0140-6736(03)13909-8
12. Lietman TM, Deiner MS, Oldenburg CE, Nash SD, Keenan JD, Porco TC. Identifying a sufficient core group for trachoma transmission. *PLoS Negl Trop Dis*. 2018;12(10):e0006478. doi:10.1371/journal.pntd.0006478
13. Chidambaram JD, Alemayehu W, Melese M, et al. Effect of a single mass antibiotic distribution on the prevalence of infectious trachoma. *JAMA*. 2006;295(10):1142-1146. doi:10.1001/jama.295.10.1142
14. Melese M, Alemayehu W, Lakew T, et al. Comparison of annual and biannual mass antibiotic administration for elimination of infectious trachoma. *JAMA*. 2008;299(7):778-784. doi:10.1001/jama.299.7.778
15. Gebre T, Ayele B, Zerihun M, et al. Comparison of annual versus twice-yearly mass azithromycin treatment for hyperendemic trachoma in Ethiopia: a cluster-randomised trial. *Lancet (London, England)*. 2012;379(9811):143-151. doi:10.1016/S0140-6736(11)61515-8
16. House JI, Ayele B, Porco TC, et al. Assessment of herd protection against trachoma due to repeated mass antibiotic distributions: a cluster-randomised trial. *Lancet*

*(London, England)*. 2009;373(9669):1111-1118. doi:10.1016/S0140-6736(09)60323-8
17. Stoller NE, Gebre T, Ayele B, et al. Efficacy of latrine promotion on emergence of infection with ocular Chlamydia trachomatis after mass antibiotic treatment: a cluster-randomized trial. *Int Health*. 2011;3(2):75-84. doi:10.1016/j.inhe.2011.03.004
18. Lietman TM, Ayele B, Gebre T, et al. Frequency of Mass Azithromycin Distribution for Ocular Chlamydia in a Trachoma Endemic Region of Ethiopia: A Cluster Randomized Trial. *Am J Ophthalmol*. 2020;214:143-150. doi:10.1016/j.ajo.2020.02.019
19. Jawetz E, Rose L, Hanna L, Thygeson P. Experimental inclusion conjunctivitis in man: measurements of infectivity and resistance. *JAMA*. 1965;194(6):620-632.
20. Liu F, Porco TC, Mkocha HA, et al. The efficacy of oral azithromycin in clearing ocular chlamydia: mathematical modeling from a community-randomized trachoma trial. *Epidemics*. 2014;6:10-17. doi:10.1016/j.epidem.2013.12.001
